# A catalog of associations between rare coding variants and COVID-19 outcomes

**DOI:** 10.1101/2020.10.28.20221804

**Authors:** J. A. Kosmicki, J. E. Horowitz, N. Banerjee, R. Lanche, A. Marcketta, E. Maxwell, X. Bai, D. Sun, J. D. Backman, D. Sharma, H. M. Kang, C. O’Dushlaine, A. Yadav, A. J. Mansfield, A. H. Li, K. Watanabe, L. Gurski, S. E. McCarthy, A. E. Locke, S. Khalid, S. O’Keeffe, J. Mbatchou, O. Chazara, Y. Huang, E. Kvikstad, A. O’Neill, P. Nioi, M. M. Parker, S. Petrovski, H. Runz, J. D. Szustakowski, Q. Wang, E. Wong, A. Cordova-Palomera, E. N. Smith, S. Szalma, X. Zheng, S. Esmaeeli, J. W. Davis, Y-P. Lai, X. Chen, A. E. Justice, J. B. Leader, T. Mirshahi, D. J. Carey, A. Verma, G. Sirugo, M. D. Ritchie, D. J. Rader, G. Povysil, D. B. Goldstein, K. Kiryluk, E. Pairo-Castineira, K. Rawlik, D. Pasko, S. Walker, A. Meynert, A. Kousathanas, L. Moutsianas, A. Tenesa, M. Caulfield, R. Scott, J. F. Wilson, J. K. Baillie, G. Butler-Laporte, T. Nakanishi, M. Lathrop, J.B. Richards, Regeneron Genetics Center, UKB Exome Sequencing Consortium, M. Jones, S. Balasubramanian, W. Salerno, A. R. Shuldiner, J. Marchini, J. D. Overton, L. Habegger, M. N. Cantor, J. G. Reid, A. Baras, G. R. Abecasis, M. A. Ferreira

## Abstract

Severe acute respiratory syndrome coronavirus-2 (SARS-CoV-2) causes coronavirus disease-19 (COVID-19), a respiratory illness that can result in hospitalization or death. We investigated associations between rare genetic variants and seven COVID-19 outcomes in 543,213 individuals, including 8,248 with COVID-19. After accounting for multiple testing, we did not identify any clear associations with rare variants either exome-wide or when specifically focusing on (i) 14 interferon pathway genes in which rare deleterious variants have been reported in severe COVID-19 patients; (ii) 167 genes located in COVID-19 GWAS risk loci; or (iii) 32 additional genes of immunologic relevance and/or therapeutic potential. Our analyses indicate there are no significant associations with rare protein-coding variants with detectable effect sizes at our current sample sizes. Analyses will be updated as additional data become available, with results publicly browsable at https://rgc-covid19.regeneron.com.

## MAIN TEXT

The severe acute respiratory syndrome coronavirus 2 (SARS-CoV-2) [1] causes coronavirus disease 2019 (COVID-19) [2]. COVID-19 ranges in clinical presentation from asymptomatic infection to flu-like illness with respiratory failure, hyperactive immune responses and death [3–5]. It is currently estimated that SARS-CoV-2 has infected >100 million individuals and has been attributed to >2 million recorded deaths worldwide. Known risk factors for severe disease include male sex, older age, ancestry, obesity and underlying cardiovascular, renal, and respiratory diseases [6–9], among others.

Since the start of the SARS-CoV-2 pandemic, host genetic analysis of common genetic variation among SARS-CoV-2 patients have identified at least nine genome-wide significant loci that modulate COVID-19 susceptibility and severity, including variants in/near *LZTFL1*, *IFNAR2*, *DPP9* and the *HLA* region [10–13]. However, to date, there has been no assessment of the contribution of rare genetic variation to COVID-19 disease susceptibility or severity through large population-based exome-wide association analyses.

To identify rare variants (RVs, minor allele frequency [MAF]<1%) associated with COVID-19 susceptibility and severity, we generated exome-wide sequencing data for 543,213 individuals from three studies (Geisinger Health System [GHS], Penn Medicine BioBank [PMBB] and UK Biobank [UKB]) and three ancestries (African, European and South Asian) (**Supplementary Table 1**). Of these, 8,248 had COVID-19, and among those 2,085 (25.28%) were hospitalized and 590 (7.15%) had severe disease (i.e. requiring ventilation or resulting in death; **Supplementary Table 2**). Using these data, we tested the association between RVs and seven COVID-19 outcomes: five related to disease susceptibility and two related to disease severity among COVID-19 cases (**Supplementary Table 3**). In a separate paper [13], we used these same phenotypes to validate the association with common risk variants reported in previous COVID-19 GWAS[10–12], thus demonstrating that our phenotypes are calibrated with those used in other studies.

For each phenotype, exome-wide association analyses were performed separately in each study and ancestry using REGENIE[14], testing single RVs (∼7 million) and a burden of RVs in 18,886 protein-coding genes. The genomic inflation factor (λ_GC_) for RVs was often <1 in individual studies, caused by a large proportion of variants having a minor allele count (MAC) of 0 in cases (**Supplementary Table 4**). In meta-analyses across studies and ancestries, we found no RV associations at a conservative *P*<9.6×10^-10^, which corresponds to a Bonferroni correction for the number of variants and traits tested. The most significant associations with RVs are listed in **Table 1**, all observed with our COVID-19 hospitalization phenotype (2,085 hospitalized cases vs. 534,965 COVID-19 negative or unknown controls). Of these, we highlight an association with an RV in the promoter of *EEF2* (rs532051930:A, MAF=0.003%, OR=93.9, 95% CI 20.3-434.5, *P*=6.2×10^-9^; **Supplementary Figure 1A**), a translation elongation factor which plays a key role in viral replication[15, 16].

**Table 1.** Top rare variant associations identified in this study (P<10^-8^), all observed with the phenotype COVID-19 positive and hospitalized (cases) vs. COVID-19 negative or unknown (controls).

| Variant | Effect allele | Odds Ratio [95% CI] | P-value | N cases with 0\|1\|2 copies of effect allele | N controls with 0\|1\|2 copies of effect allele | Effect allele frequency | Gene | Variant effect |
| --- | --- | --- | --- | --- | --- | --- | --- | --- |
| rs374698271 | T | 81.60 [18.71,355.81] | 4.65E-09 | 1698\|5\|0 | 399222\|18\|0 | 0.00003 | *RFX2* | Intronic |
| rs532051930 | A | 93.90 [20.29,434.50] | 6.17E-09 | 1869\|4\|0 | 508510\|22\|0 | 0.00003 | *EEF2* | 5 prime UTR |
| rs751932982 | A | 22.68 [7.89,65.23] | 6.92E-09 | 1869\|2\|2 | 508247\|44\|8 | 0.00006 | *FAM9B* | Intronic |

Despite not reaching our threshold for genome-wide significance, we highlight the association with *EEF2* because: (i) it was supported by two independent studies (**Supplementary Figure 1B**); (ii) five additional RVs in the 89 bp promoter of *EEF2* (out of 50 tested) had an independent and directionally-consistent predisposing association with the same hospitalization phenotype (*P*<0.05; **Supplementary Table 5**); (iii) rs532051930 is predicted by DeepSEA [17] to be a regulatory variant and to disrupt binding of several transcription factors, including GABP, which has been linked to RV-induced aberrant gene expression [18]; (iv) rs532051930 is located at the peak of the transcription initiation region for *EEF2* (**Supplementary Figure 1C**) [19], suggesting that it might affect RNA polymerase II binding; and (v) among a subset of 1,988 individuals with available RNA-seq data from liver tissue, the only individual who was a carrier for rs532051930:A had the second highest expression of *EEF2* (**Supplementary Figure 1D**). These results raise the possibility that rs532051930 and potentially other promoter RVs increase *EEF2* transcription and, consequently, increase risk of hospitalization due to SARS-CoV-2 infection.

Given these supporting observations and the established role of *EEF2* in viral replication, we studied the genetic association with rs532051930 in greater detail, to understand if it was likely to be a true-positive association. First, we reviewed sequencing reads for all 26 carriers of this variant, to visually validate the heterozygote genotype call produced by the calling algorithm. Sequencing reads were consistent with a heterozygote call for all 26 individuals. Second, we determined if the association with this rare variant was robust to the association test used (**Supplementary Table 6**). A Firth test applied to a joint analysis of data across the two studies, adjusting for study specific covariates as an offset, resulted in *P*=5.5×10^-8^. Similar results were obtained by: (i) an approximation to this approach, which combines log-likelihood curves calculated at a grid of values with the Firth penalty, applied using approximate derivatives; (ii) a *P*-value-based meta-analysis, for which there is evidence of better type-I error control [20]; and (iii) the BinomiRare test [21], which uses a test statistic based on allele counts in cases only, adjusts for covariates and combines data across studies. We also applied tests without covariates and these uniformly led to less significant associations, but this is expected since the covariates explain variation in the phenotype. Third, we attempted to replicate this association by querying whether this variant was present in an additional 4,341 COVID-19 cases with exome- or whole-genome sequence data generated as part of the GenOMICC (n=2,969) [11], Columbia University COVID-19 Biobank (n=1,152) and Biobanque Quebec (n=220) [22] studies. We found no carriers for this variant in these additional COVID-19 cases (**Supplementary Table 7**). Given these findings, we conclude that it is not certain that there is a true association between rs532051930 and COVID-19 risk, illustrating the importance of replication.

Next, we addressed the possibility that associations with protein-coding RVs might help pinpoint target genes of common risk variants identified in GWAS of COVID-19. To this end, we focused on 167 genes located within 500 kb of the nine common variants associated with COVID-19 hospitalization in a separate analysis [13]. Of the 334 gene burden tests performed (167 genes x 2 burden tests), which considered both pLoF variants alone (M1 burden test) or pLoF plus deleterious missense variants (M3 burden test), 17 had a nominally significant association with the same COVID-19 hospitalization phenotype (**Supplementary Table 6**). Of these, burden tests for one gene – *CHAF1A* (OR=25 for the M1 burden test, 95% CI 4.9-128.5, *P*=1.0×10^-4^) *–* remained borderline significant after correcting for the 334 tests performed (*P*<0.05/334=1.5×10^-4^). *CHAF1A* is located 317 kb from the lead common variant at locus 19p13.3 [13], is highly expressed in EBV-transformed B-cells [23] and encodes a component of the chromatin assembling factor complex that affects cell differentiation, including the differentiation of pre-B cells into B-cells and macrophages [24].

We then examined the association with 14 genes in the interferon pathway, given recent reports that deleterious RVs in these genes may be implicated in severe clinical outcomes [25, 26]. Given the larger sample size in our studies, we examined whether there was any evidence for association between the COVID-19 hospitalization phenotype (2,085 cases vs. 534,965 controls) and the burden of rare (MAF<0.1%) pLoF variants (M1 burden test) or pLoF plus deleterious missense variants (M3 burden test) in these 14 genes, three of which are located within 500 kb of a COVID-19 GWAS risk variant (*IFNAR1*, *IFNAR2* and *TICAM1*). Of the 14 genes, only two genes had a nominal significant association: *STAT2* and *TLR7* (**Table 2**). However, neither remained significant after correcting for the 28 tests performed (both with *P*>0.05/28=0.0018). Further, these results were unchanged when testing COVID-19 severe cases (N=590), or when restricting the burden tests to include variants with a MAF<1% or singleton variants (**Supplementary Table 7**). Therefore, as recently reported by others [22], we found no evidence for an association between RVs in these 14 interferon signaling genes and risk of COVID-19.

**Table 2.** Association between the phenotype COVID-19 positive and hospitalized (cases) vs COVID-19 negative or unknown (controls) and 14 genes related to interferon signaling that were recently reported to contain rare (MAF<0.1%), deleterious variants in patients with severe COVID-19 [14, 23].

| Variants included in burden test | Gene | Odds Ratio (95% CI) | P-value | N cases with RR\|RA\|AA genotype* | N controls with RR\|RA\|AA genotype* | AAF |
| --- | --- | --- | --- | --- | --- | --- |
| pLoF, MAF<0.1% | *IFNAR1* | 0.805[0.146;4.438] | 8.00E-01 | 2019\|1\|0 | 525406\|359\|0 | 0.00034 |
|  | *IFNAR2* | 2.077[0.620;6.961] | 2.40E-01 | 2082\|3\|0 | 534277\|688\|0 | 0.00064 |
|  | *IKBKG* | 0.491[0.005;50.219] | 7.60E-01 | 1873\|0\|0 | 508491\|31\|10 | 0.00005 |
|  | *IRF3* | 0.956[0.251;3.643] | 9.50E-01 | 2083\|2\|0 | 534559\|406\|0 | 0.00038 |
|  | *IRF7* | 1.165[0.426;3.185] | 7.70E-01 | 2082\|3\|0 | 534124\|841\|0 | 0.00079 |
|  | *IRF9* | 0.371[0.003;53.766] | 7.00E-01 | 1873\|0\|0 | 508479\|53\|0 | 0.00005 |
|  | *STAT1* | 0.365[0.001;126.712] | 7.40E-01 | 1873\|0\|0 | 508490\|42\|0 | 0.00004 |
|  | *STAT2* | 0.355[0.028;4.438] | 4.20E-01 | 1873\|0\|0 | 508405\|127\|0 | 0.00012 |
|  | *TBK1* | 0.365[0.011;11.964] | 5.70E-01 | 1873\|0\|0 | 508445\|87\|0 | 0.00009 |
|  | *TICAM1* | 3.730[0.189;73.610] | 3.90E-01 | 1872\|1\|0 | 508368\|164\|0 | 0.00016 |
|  | *TLR3* | 1.128[0.144;8.834] | 9.10E-01 | 2084\|1\|0 | 534674\|291\|0 | 0.00027 |
|  | *TLR7* | 7.627[1.872;31.075] | 4.60E-03 | 1872\|0\|1 | 508503\|25\|4 | 0.00003 |
|  | *TRAF3* | 0.368[0.000;733.638] | 8.00E-01 | 1873\|0\|0 | 508504\|28\|0 | 0.00003 |
|  | *UNC93B1* | 1.302[0.272;6.238] | 7.40E-01 | 1938\|2\|0 | 516861\|409\|0 | 0.0004 |
| pLoF or missense predicted deleterious, MAF<0.1% | *IFNAR1* | 0.756[0.234;2.444] | 6.40E-01 | 2083\|2\|0 | 534219\|746\|0 | 0.0007 |
|  | *IFNAR2* | 1.968[0.598;6.469] | 2.70E-01 | 2082\|3\|0 | 534253\|712\|0 | 0.00067 |
|  | *IKBKG* | 0.446[0.010;19.737] | 6.80E-01 | 1873\|0\|0 | 508452\|70\|10 | 0.00009 |
|  | *IRF3* | 0.786[0.238;2.592] | 6.90E-01 | 2083\|2\|0 | 534413\|552\|0 | 0.00052 |
|  | *IRF7* | 1.137[0.519;2.492] | 7.50E-01 | 2080\|5\|0 | 533486\|1479\|0 | 0.00138 |
|  | *IRF9* | 0.371[0.003;53.766] | 7.00E-01 | 1873\|0\|0 | 508479\|53\|0 | 0.00005 |
|  | *STAT1* | 0.365[0.038;3.488] | 3.80E-01 | 2018\|0\|0 | 526009\|218\|0 | 0.00021 |
|  | *STAT2* | 2.600[1.272;5.314] | 8.80E-03 | 2073\|12\|0 | 533405\|1559\|1 | 0.00146 |
|  | *TBK1* | 1.114[0.445;2.790] | 8.20E-01 | 2081\|4\|0 | 533861\|1103\|1 | 0.00103 |
|  | *TICAM1* | 3.657[0.188;71.218] | 3.90E-01 | 1872\|1\|0 | 508365\|167\|0 | 0.00016 |
|  | *TLR3* | 0.805[0.435;1.493] | 4.90E-01 | 2077\|8\|0 | 532355\|2609\|1 | 0.00244 |
|  | *TLR7* | 1.580[0.627;3.979] | 3.30E-01 | 2001\|3\|1 | 525830\|477\|163 | 0.00076 |
|  | *TRAF3* | 3.013[0.527;17.217] | 2.10E-01 | 2019\|1\|0 | 525523\|242\|0 | 0.00023 |
|  | *UNC93B1* | 1.665[0.774;3.582] | 1.90E-01 | 2075\|10\|0 | 533331\|1634\|0 | 0.00153 |
| * RR: individuals who have genotype Reference/Reference for all variants included in burden test. RA: individuals who have genotype Reference/Alternate for at least one variant. AA: individuals who have genotype Alternate/Alternate for at least one variant. |  |  |  |  |  |  |

Lastly, we performed the same analysis for an additional 32 genes that are involved in the etiology of SARS-CoV-2 infection (*ACE2, TMPRSS2*), encode therapeutic targets for COVID-19 obtained through ClinicalTrials.gov (*e.g. IL6R, JAK1*) or have been implicated in other immune or infectious diseases through GWAS (*e.g. IL33*). After correcting for multiple testing, there were also no significant associations with a burden of deleterious RVs for this group of COVID-19 therapeutic target genes (**Supplementary Table 8**).

In summary, we provide a catalog of RV associations with COVID-19 outcomes based on exome-sequence data, capturing genetic variation not assayed by array genotyping or imputation. We did not find any convincing associations with current sample sizes, but will continue to expand our analyses and update results periodically at https://rgc-covid19.regeneron.com.

## METHODS

### Participating Studies

#### Geisinger Health System (GHS)

The GHS MyCode Community Health Initiative study has been described previously [27]. Briefly, the GHS study is a health system-based cohort from central and eastern Pennsylvania (USA) with ongoing recruitment since 2006. A subset of 144,182 MyCode participants sequenced as part of the GHS-Regeneron Genetics Center DiscovEHR partnership were included in this study. Information on COVID-19 outcomes were obtained through GHS’s COVID-19 registry. Patients were identified as eligible for the registry based on relevant lab results and ICD-10 diagnosis codes; patient charts were then reviewed to confirm COVID-19 diagnoses. The registry contains data on outcomes, comorbidities, medications, supplemental oxygen use and ICU admissions.

#### Penn Medicine BioBank (PMBB) study

PMBB study participants are recruited through the University of Pennsylvania Health System, which enrolls participants during hospital or clinic visits. Participants donate blood or tissue and allow access to EHR information[28]. The PMBB COVID-19 registry consists of patients who have positive qPCR testing for SARS-COV-2. We then used electronic health records to classify COVID-19 patients into hospitalized and severe (ventilation or death) categories.

#### UK Biobank (UKB) study

We studied the host genetics of SARS-CoV-2 infection in participants of the UK Biobank study, which took place between 2006 and 2010 and includes approximately 500,000 adults aged 40-69 at recruitment. In collaboration with UK health authorities, the UK Biobank has made available regular updates on COVID-19 status for all participants, including results from four main data types: qPCR test for SARS-CoV-2, anonymized electronic health records, primary care and death registry data. We report results based on the 12 September 2020 data refresh and excluded from the analysis 28,547 individuals with a death registry event prior to 2020.

### COVID-19 phenotypes used for genetic association analyses

We grouped participants from each study into three broad COVID-19 disease categories (**Supplementary Table 2**): (i) positive – those with a positive qPCR or serology test for SARS-CoV-2, or a COVID-19-related ICD10 code (U07), hospitalization or death; (ii) negative – those with only negative qPCR or serology test results for SARS-CoV-2 and no COVID-19-related ICD10 code (U07), hospitalization or death; and (iii) unknown – those with no qPCR or serology test results and no COVID-19-related ICD10 code (U07), hospitalization or death. We then used these broad COVID-19 disease categories, in addition to hospitalization and disease severity information, to create seven COVID-19-related phenotypes for genetic association analyses, as detailed in **Supplementary Table 3**.

### Array genotyping

Genotyping was performed on one of four SNP array types: Illumina OmniExpress Exome array (OMNI; 59345 samples from GHS), Illumina Global Screening Array (GSA; PMBB and 82,527 samples from GHS), Applied Biosystems UK BiLEVE Axiom Array (49,950 samples from UKB), or Applied Biosystems UK Biobank Axiom Array (438,427 samples from UKB). We retained variants with a minor allele frequency (MAF) >1%, <10% missingness, Hardy-Weinberg equilibrium test *P*-value>10^-15^. Array data were then used: (i) to define ancestry subsets; and (ii) to generate a polygenic risk score (PRS) predictor, as part of the exome-wide association analyses carried out in REGENIE (see below).

### Exome sequencing

#### Sample Preparation and Sequencing

Genomic DNA samples normalized to approximately 16 ng/ul were transferred to the Regeneron Genetics Center from the UK Biobank in 0.5ml 2D matrix tubes (Thermo Fisher Scientific) and stored in an automated sample biobank (LiCONiC Instruments) at −80°C prior to sample preparation. Exome capture was completed using a high-throughput, fully-automated approach developed at the Regeneron Genetics Center. Briefly, DNA libraries were created by enzymatically shearing 100ng of genomic DNA to a mean fragment size of 200 base pairs using a custom NEBNext Ultra II FS DNA library prep kit (New England Biolabs) and a common Y-shaped adapter (Integrated DNA Technologies [IDT]) was ligated to all DNA libraries. Unique, asymmetric 10 base pair barcodes were added to the DNA fragment during library amplification with KAPA HiFi polymerase (KAPA Biosystems) to facilitate multiplexed exome capture and sequencing. Equal amounts of sample were pooled prior to overnight exome capture, approximately 16 hours, with either (i) a slightly modified version of IDT’s xGen probe library (for UKB, PMBB and 81,620 samples of GHS); or (ii) NimbleGen VCRome (58,856 samples of GHS). Captured fragments were bound to streptavidin-coupled Dynabeads (Thermo Fisher Scientific) and non-specific DNA fragments removed through a series of stringent washes using the xGen Hybridization and Wash kit according to the manufacturer’s recommended protocol (Integrated DNA Technologies). The captured DNA was PCR amplified with KAPA HiFi and quantified by qPCR with a KAPA Library Quantification Kit (KAPA Biosystems). The multiplexed samples were pooled and then sequenced using: (i) for UKB samples – 75 bp paired-end reads with two 10 base pair index reads on the Illumina NovaSeq 6000 platform using S2 or S4 flow cells; (ii) for GHS samples captured with VCRome – 75 bp paired-end reads with two 8 bp index reads on the Illumina HiSeq 2500; (iii) for GHS captured with IDT – two 8 bp index reads on the Illumina HiSeq 2500 or two 10 bp index reads on the Illumina NovaSeq 6000 on S4 flow cells; (iv) for UPENN-PMBB – two 10 bp index reads on the Illumina NovaSeq 6000 on S4 flow cells.

#### Variant calling and quality control

Sample read mapping and variant calling, aggregation and quality control were performed via the SPB protocol described in Van Hout et al. [29]. Briefly, for each sample, NovaSeq WES reads are mapped with BWA MEM to the hg38 reference genome. Small variants are identified with WeCall and reported as per-sample gVCFs. These gVCFs are aggregated with GLnexus into a joint-genotyped, multi-sample VCF (pVCF). SNV genotypes with read depth (DP) less than seven and indel genotypes with read depth less than ten are changed to no-call genotypes. After the application of the DP genotype filter, a variant-level allele balance filter is applied, retaining only variants that meet either of the following criteria: (i) at least one homozygous variant carrier or (ii) at least one heterozygous variant carrier with an allele balance (AB) greater than the cutoff (AB ≥ 0.15 for SNVs and AB ≥ 0.20 for indels).

#### Identification of low-quality variants from exome-sequencing using machine learning

Briefly, in each study, we defined a set of positive control and negative control variants based on: (i) concordance in genotype calls between array and exome sequencing data; (ii) Mendelian inconsistencies in the exome sequencing data; (iii) differences in allele frequencies between exome sequencing batches (UKB and GHS); (iv) variant loadings on 20 principal components derived from the analysis of variants with a MAF<1%; (v) transmitted singletons. The model was then trained on up to 30 available WeCall/GLnexus site quality metrics, including, for example, allele balance and depth of coverage. We split the data into training (80%) and test (20%) sets. We performed a grid search with 5-fold cross-validation on the training set to identify the hyperparameters that return the highest accuracy during cross-validation, which are then applied to the test set to confirm accuracy. This approach identified as low-quality a total of 7 million variants in the UKB study (86% in the buffer region), 7.2 million across the two GHS datasets (IDT and VCRome; 84% in the buffer region) and 1.1 million in the PMBB study (88% in the buffer region). These variants were removed from analysis in the respective studies.

#### Gene burden masks

Briefly, for each gene region as defined by Ensembl [30], genotype information from multiple rare coding variants was collapsed into a single burden genotype, such that individuals who were: (i) homozygous reference (Ref) for all variants in that gene were considered homozygous (RefRef); (ii) heterozygous for at least one variant in that gene were considered heterozygous (RefAlt); (iii) and only individuals that carried two copies of the alternative allele (Alt) of the same variant were considered homozygous for the alternative allele (AltAlt). We did not phase rare variants; compound heterozygotes, if present, were considered heterozygous (RefAlt). We did this separately for four classes of variants: (i) predicted loss of function (pLoF), which we refer to as an “M1” burden mask; (ii) pLoF or missense (“M2”); (iii) pLoF or missense variants predicted to be deleterious by 5/5 prediction algorithms (“M3”); (iv) pLoF or missense variants predicted to be deleterious by 1/5 prediction algorithms (“M4”). Variants were annotated using SnpEff 4.3[31] and the most severe consequence for each variant was chosen, considering complete protein-coding transcripts for each gene. The following variants were considered to be pLoF variants: frameshift-causing indels, variants affecting splice acceptor and donor sites, variants leading to stop gain, stop loss and start loss. The five missense deleterious algorithms used were SIFT [32], PolyPhen2 (HDIV), PolyPhen2 (HVAR) [33], LRT [34], and MutationTaster [35]. For each gene, and for each of these four groups, we considered five separate burden masks, based on the frequency of the alternative allele of the variants that were screened in that group: <1%, <0.1%, <0.01%, <0.001% and singletons only. Each burden mask was then tested for association with the same approach used for individual variants (see below).

### Genetic association analyses

Association analyses in each study were performed using the genome-wide Firth logistic regression test implemented in REGENIE [14]. In this implementation, Firth’s approach is applied when the p-value from standard logistic regression score test is below 0.05. As the Firth penalty (*i.e.* Jeffrey’s invariant prior) corresponds to a data augmentation procedure where each observation is split into a case and a control with different weights, it can handle variants with no minor alleles among cases. With no covariates, this corresponds to adding 0.5 in every cell of a 2×2 table of allele counts versus case-control status.

We included in step 1 of REGENIE (*i.e.* prediction of individual trait values based on the genetic data) array variants with a minor allele frequency (MAF) >1%, <10% missingness, Hardy-Weinberg equilibrium test *P*-value>10^-15^ and linkage-disequilibrium (LD) pruning (1000 variant windows, 100 variant sliding windows and *r*^2^<0.9). The exception was the GHS study, for which we used exome (not array) variants in step 1; we did this because two different exome capture technologies (IDT and VCRome) were used to sequence the GHS samples, and so it was important to capture in step 1 of REGENIE any differences in exome sequencing performance between IDT and VCRome. We excluded from step 1 any SNPs with high inter-chromosomal LD, in the major histo-compatibility (MHC) region, or in regions of low complexity.

The association model used in step 2 of REGENIE included as covariates (i) age, age^2^, sex, age-by-sex and age^2^-by-sex; (ii) 10 ancestry-informative principle components (PCs) derived from the analysis of a set of LD-pruned (50 variant windows, 5 variant sliding windows and *r*^2^<0.5) common variants from the array (imputed for the GHS study) data; (iii) an indicator for exome sequencing batch (GHS: two IDT batches, one VCRome batch; UKB: six IDT batches); and (iv) 20 PCs derived from the analysis of exome variants with a MAF between 2.6×10^-5^ (roughly corresponding to a minor allele count [MAC] of 20) and 1%. We corrected for PCs built from rare variants because previous studies demonstrated PCs derived from common variants do not adequately correct for fine-scale population structure [36, 37].

Within each study, association analyses were performed separately for five different continental ancestries defined based on the array data: African (AFR), Admixed American (AMR), European (EUR) and South Asian (SAS). We determined continental ancestries by projecting each sample onto reference principle components calculated from the HapMap3 reference panel. Briefly, we merged our samples with HapMap3 samples and kept only SNPs in common between the two datasets. We further excluded SNPs with MAF<10%, genotype missingness >5% or Hardy-Weinberg Equilibrium test p-value < 10^-5^. We calculated PCs for the HapMap3 samples and projected each of our samples onto those PCs. To assign a continental ancestry group to each non-HapMap3 sample, we trained a kernel density estimator (KDE) using the HapMap3 PCs and used the KDEs to calculate the likelihood of a given sample belonging to each of the five continental ancestry groups. When the likelihood for a given ancestry group was >0.3, the sample was assigned to that ancestry group. When two ancestry groups had a likelihood >0.3, we arbitrarily assigned AFR over EUR, AMR over EUR, AMR over EAS, SAS over EUR, and AMR over AFR. Samples were excluded from analysis if no ancestry likelihoods were >0.3, or if more than three ancestry likelihoods were > 0.3.

Results were subsequently meta-analyzed across studies and ancestries using an inverse variance-weighed fixed-effects meta-analysis.

### Gene expression analysis in participants of the GHS study

For a subset of individuals from the GHS study (n=1,988, ascertained through the Geisinger Bariatric Surgery Clinic), RNA was extracted from liver biopsies conducted during bariatric surgery to evaluate liver disease. Individuals had class 3 obesity (BMI>40kg/m^2^) or class 2 obesity (BMI 35-39 kg/m^2^) with an obesity-related co-morbidity (e.g. type-2 diabetes, hypertension, sleep apnea, non-alcoholic fatty liver disease). RNA libraries were prepared using polyA-extraction and then sequenced with 75bp paired-end reads with two 10 bp index reads on the Illumina NovaSeq 6000 on S4 flow cells. RNA-seq data were then analyzed using the GTEx v8 workflow[38], using STAR [39] and RNASeqQC [40], except that GENCODE v32 was used in lieu of v26. Briefly: (i) raw expression counts were normalized with TMM (Trimmed Mean of M-values) as implemented in edgeR [40]; (ii) a rank-based inverse normal transformation was applied to the normalized expression values; (iii) principal components (PCs) analysis was performed on data from 25,078 genes with TPM >0.1 in >20% samples, to identify latent factors accounting for variation in gene expression; (iv) gene expression levels were adjusted for the top 100 PCs to improve power to identify cis-regulatory effects.

### Frequency of *EEF2* rare promoter variant in COVID-19 cases from independent studies

To help understand if the association between COVID-19 risk and rs532051930 in *EEF2* was likely to be a true-positive association, we determine its frequency in 4,341 cases from three additional studies.

#### GenOMICC (n=2,969)

Individuals with severe COVID-19 were ascertained as described previously[11]. DNA samples were then whole-genome sequenced on the Illumina NovaSeq 6000 platform, aligned to the human reference genome hg38 and variant called to GVCF stage on the DRAGEN pipeline (software v01.011.269.3.2.22, hardware v01.011.269) at Genomics England. rs532051930 +/-50bp was genotyped with the GATK GenotypeGVCFs tool v4.1.8.1 and filtered to minimum depth 8X. Ancestry for individuals with array genotyping was inferred using ADMIXTURE[41] populations defined in 1000 Genomes[42]. When one individual had a probability > 80% of pertaining to one ancestry, then the individual was assigned to this ancestry, otherwise the individual was considered to be of admixed ancestry, as performed in the Million veteran program [43]. Somalier v0.2.12[44] was used to estimate ancestry for samples with whole-genome sequencing data. Predictions from Somalier were compared against predictions from ADMIXTURE for 1833 samples with both array genotyping and whole-genome sequencing data. Of these, the ancestry assignment matched between the two approaches for 1832 samples. Somalier predictions were used for the remaining 927 samples, of which 813 could be confidently (≥95% probability) assigned to a population; the remaining 114 were assigned to admixed ancestry.

#### Columbia University COVID-19 biobank (n=1,152)

This cohort has previously been described in detail[22]. Briefly, 1,152 COVID-19 patients that were treated for COVID-19 at the Columbia University Irving Medical Center were recruited to the Columbia University COVID-19 Biobank between March and May 2020. All patients had PCR-confirmed SARS-CoV-2 infection and the vast majority had severe COVID-19 requiring hospitalization. For all cases, exomes were captured with the IDT xGen Exome Research Panel V1.0 and sequenced on Illumina’s NovaSeq 6000 platform with 150 bp paired-end reads according to standard protocols. All cases were processed with the same bioinformatic pipeline for variant calling. In brief, reads were aligned to human reference GRCh37 using DRAGEN and duplicates were marked with Picard. Variants were called according to the Genome Analysis Toolkit (GATK) Best Practices recommendations v3.66[45]. Finally, variants were annotated with ClinEff[31] and the IGM’s in-house tool ATAV[46]. A centralized database was used to store variant and per site coverage data for all samples enabling well controlled analyses without the need of generating jointly called VCF files (see Ren et al. 2020 for details[46]). For each patient, we performed ancestry classification into one of the six major ancestry groups (European, African, Latinx, East Asian, South Asian and Middle Eastern) using a neural network trained on a set of samples with known ancestry labels. We used a 50% probability cut-off to assign an ancestry label to each sample and labeled samples that did not reach 50% for any of the ancestral groups as “Admixed”.

We only included samples that had at least 90% of the consensus coding sequence (CCDS release 20[47]) covered at ≥ 10x and ≤ 3% contamination levels according to VerifyBamID[48]. Additionally, we removed samples with a discordance between self-declared and sequence-derived gender and samples with an inferred relationship of second-degree or closer according to KING[49]. All cases had at least 10x coverage at the position of rs532051930.

#### Biobanque Québec Covid-19 (n=220)

The Biobanque Québec COVID-19 (www.BQC19.ca) is a provincial biobank prospectively enrolling patients with suspected COVID-19, or COVID-19 confirmed through SARS-CoV-2 PCR testing and was previously described[22]. For this study, we used results from patients with available WGS data and who were recruited at the Jewish General Hospital (JGH) in Montreal. The JGH is a university affiliated hospital serving a large multi-ethnic adult population and the Québec government designated the JGH as the primary COVID-19 reference center early in the pandemic. In total, Biobanque Quebec contained 533 participants with WGS, including 62 cases of COVID-19 who required invasive ventilatory support (BiPAP, high flow oxygen, or endotracheal intubation) or died, 128 COVID-19 patients who were hospitalized but did not require invasive ventilatory support, 30 individuals with COVID-19 did not require hospitalization, and 313 SARS-CoV-2 PCR-negative participants. Using genetic PCAs derived from genome-wide genotyping, 76% of participants were of European ancestry, 9% were of African ancestry, 7% were of east Asian ancestry, and 5% were of south Asian ancestry.

We performed WGS at a mean depth of 30x on all individuals using Illumina’s Novaseq 6000 platform (Illumina, San Diego, CA, USA). Sequencing results were analyzed using the McGill Genome Center bioinformatics pipelines[50], in accordance with Genome Analysis Toolkit (GATK) best practices recommendations[45]. Reads were aligned to the GRCh38 reference genome. Variant quality control was performed using the variantRecalibrator and applyVQSR functions from GATK.

## Results availability

All genotype-phenotype association results reported in this study are available for browsing using the RGC’s COVID-19 Results Browser (https://rgc-covid19.regeneron.com). Data access and use is limited to research purposes in accordance with the Terms of Use (https://rgc-covid19.regeneron.com/terms-of-use). The COVID-19 Results Browser provides a user-friendly interface to explore genetic association results, enabling users to query summary statistics across multiple cohorts and association studies using genes, variants or phenotypes of interest. Results are displayed in an interactive tabular view ordered by p-value – enabling filtering, sorting, grouping and viewing additional statistics – with link outs to individual GWAS reports, including interactive Manhattan and QQ plots. LocusZoom views of LD information surrounding variants of interest are also available, with LD calculated using the respective source genetic datasets.

The data resource supporting the COVID-19 Results Browser was built using a processed version of the raw association analysis outputs. Using the RGC’s data engineering toolkit based in Apache Spark and Project Glow (https://projectglow.io/), association results were annotated, enriched and partitioned into a distributed, columnar data store using Apache Parquet. Processed Parquet files were registered with AWS Athena, which enables efficient, scalable queries on unfiltered association result datasets. Additionally, “filtered” views of associations significant at a threshold of p-value < 0.001 were stored in AWS RDS Aurora databases for low latency queries to service primary views of top associations. APIs into RDS and Athena are managed behind the scenes such that results with a p-value>0.001 are pulled from Athena as needed.

## Data Availability

Association results reported in this manuscript are publicly available at https://rgc-covid19.regeneron.com

## Acknowledgements

This research has been conducted using the UK Biobank Resource (Project 26041). The Penn Medicine BioBank is funded by a gift from the Smilow family, the National Center for Advancing Translational Sciences of the National Institutes of Health under CTSA Award Number UL1TR001878, and the Perelman School of Medicine at the University of Pennsylvania. Whole genome sequencing of the Biobanque Québec Covid-19 cohort was funded by the CanCOGeN HostSeq project. The Richards research group is supported by the Canadian Institutes of Health Research (CIHR), the Lady Davis Institute of the Jewish General Hospital, the Canadian Foundation for Innovation, the NIH Foundation, Cancer Research UK and the Fonds de Recherche Québec Santé (FRQS). G.B.L. is supported by a joint research fellowship from Quebec’s ministry of health and social services, and the FRQS. T.N. is supported by Research Fellowships of Japan Society for the Promotion of Science (JSPS) for Young Scientists and JSPS Overseas Challenge Program for Young Researchers. J.B.R. is supported by a FRQS Clinical Research Scholarship. The Columbia University Biobank was supported by Columbia University and the National Center for Advancing Translational Sciences, NIH, through Grant Number UL1TR001873. Columbia University COVID-19 Biobank members that additionally contributed to this work include Muredach P. Reilly, Wendy Chung, Eldad Hod, Soumitra Sengupta, Danielle Pendrick, Nitin Bhardwaj, Ning Shang, Atlas Khan, Chen Wang, Sheila M. O’Byrne, Renu Nandakumar, Amritha Menon, Yat S. So, Richard Mayeux, Ali G. Gharavi, Iuliana Ionita-Laza, Andrea Califano, Christine K. Garcia, Peter Sims, and Anne-Catrin Uhlemann. The content is solely the responsibility of the authors and does not necessarily represent the official views of the NIH or Columbia University. GenOMICC was funded by Sepsis Research (the Fiona Elizabeth Agnew Trust), the Intensive Care Society, a Wellcome-Beit Prize award to J. K. Baillie (Wellcome Trust 103258/Z/13/A), a BBSRC Institute Program Support Grant to the Roslin Institute (BBS/E/D/20002172, BBS/E/D/10002070 and BBS/E/D/30002275), the Medical Research Council [grant MC_PC_19059]. Research performed at the Human Genetics Unit was funded by the MRC (MC_UU_00007/10, MC_UU_00007/15). Whole-genome sequencing was done in partnership with Genomics England and was funded by UK Department of Health and Social Care, UKRI and LifeArc. Genomics England and the 100,000 Genomes Project was funded by the National Institute for Health Research, the Wellcome Trust, the Medical Research Council, Cancer Research UK, the Department of Health and Social Care and NHS England. M Caulfield is an NIHR Senior Investigator. This work is part of the portfolio of translational research at the NIHR Biomedical Research Centre at Barts and Cambridge. LK was supported by an RCUK Innovation Fellowship from the National Productivity Investment Fund (MR/R026408/1). We acknowledge support from the MRC Human Genetics Unit programme grant, “Quantitative traits in health and disease” (U. MC_UU_00007/10). A. Tenesa acknowledges funding from MRC research grant MR/P015514/1, and HDR-UK award HDR-9004 and HDR-9003. Recruitment to GenOMICC was enabled by the National Institute of Healthcare Research Clinical Research Network (NIHR CRN) and the Chief Scientist Office (Scotland), who facilitate recruitment into research studies in NHS hospitals, and to the global ISARIC and InFACT consortia. We thank the patients and their loved ones who volunteered to contribute to this study at one of the most difficult times in their lives, and the research staff in every intensive care unit who recruited patients at personal risk during the most extreme conditions we have ever witnessed in UK hospitals.

## Supplementary Figures

**Supplementary Figure 1.**
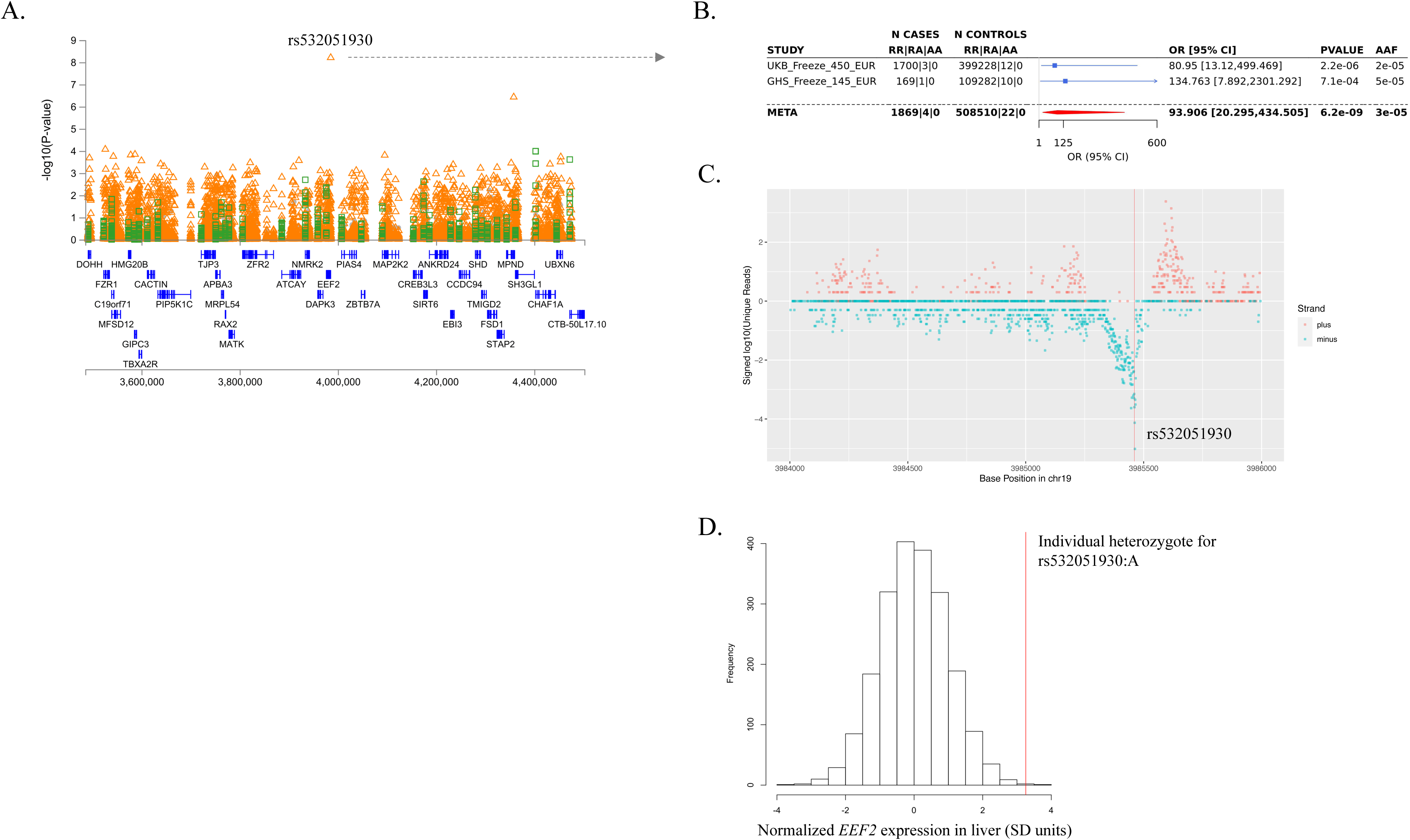
Association between a rare promoter variant in EEF2 (rs532051930:A) and the COVID-19 hospitalization phenotype. **(A)** Regional association plot centered on rs532051930. Orange triangles: individual rare variants (MAF<0.5%). Green squares: burden tests. Grey circles: individual common variants (MAF>0.5%). **(B)** Forest plot showing association in the two individual datasets included in the meta-analysis of this variant. **(C)** Results from a population-scale PROcap (Precision Run-On 5’ cap sequencing) study [19], which profiles transcription start sites of nascent RNAs attached to RNA polymerase. The variant rs532051930 (vertical red line) is located 2 bp away from the peak of transcription initiation. (**D**) Distribution of *EEF2* expression among 1,918 individuals from the GHS study with available RNA-seq data from liver tissue. Vertical red line indicates expression levels for the only individual who was heterozygote for the rare promoter variant rs532051930.

## SUPPLEMENTARY TABLES

**Supplementary Table 1.** Demographics and clinical characteristics of study participants.

| Demographics | COVID-19 Positive |  | Covid-19 Negative or Unknown |
| --- | --- | --- | --- |
|  | Hospitalized | Not Hospitalized |  |
| UK Biobank |  |  |  |
| Total N | 1848 | 5400 | 416935 |
| AFR ancestry, n (%) | 80 (3.8) | 134 (2.2) | 8495 (1.8) |
| EUR ancestry, n (%) | 1703 (81.9) | 5030 (83.2) | 399240 (85.7) |
| SAS ancestry, n (%) | 65 (3.1) | 236 (3.9) | 9200 (1.9) |
| Average Age, y (% >60y) | 59.7 (57) | 52.6 (22) | 56.2 (36) |
| Female, n (%) | 737 (39.8) | 2896 (53.6) | 229572 (55.1) |
| Hypertension, n (%) | 884 (47.8) | 1214 (22.5) | 96851 (23.2) |
| Cardiovascular Disease, n (%) | 265 (14.3) | 330 (6.1) | 24859 (5.8) |
| Type 2 Diabetes, n (%) | 342 (18.5) | 378 (7) | 25335 (6.1) |
| Chronic kidney disease, n (%) | 134 (7.3) | 112 (2.1) | 8096 (1.9) |
| Asthma, n (%) | 343 (18.5) | 886 (16.4) | 59229 (14.2) |
| COPD, n (%) | 208 (11.2) | 164 (3) | 10117 (2.4) |
| GHS |  |  |  |
| Total N | 170 | 664 | 109292 |
| EUR ancestry, n (%) | 170 (100) | 664 (100) | 109292 (100) |
| Average Age, y (% >60y) | 67.81 (76.47) | 54.74 (39.0) | 55.75 (44.14) |
| Female, n (%) | 90 (52.94) | 435 (65.51) | 68226 (62.42) |
| Hypertension, n (%) | 138 (81.17) | 343 (51.65) | 53658 (49.09) |
| Cardiovascular Disease, n (%) | 77 (45.29) | 108 (16.26) | 17269 (15.80) |
| Type 2 Diabetes, n (%) | 73 (42.94) | 158 (23.79) | 23290 (21.30) |
| Chronic kidney disease, n (%) | 64 (37.64) | 94 (14.15) | 14246 (13.03) |
| Asthma, n (%) | 15 (8.82) | 47 (7.07) | 6975 (6.38) |
| COPD, n (%) | 47(27.64) | 67 (10.09) | 10822 (9.90) |
| PMBB |  |  |  |
| Total N | 67 | 99 | 959 |
| AFR ancestry, n (%) | 67 (100) | 99 (100) | 959 (100) |
| Average Age, y (% >60y) | 62.58 (61.19) | 47.64 (19.19) | 56.24 (43.27) |
| Female, n (%) | 38 (56.7) | 72 (72.7) | 641 (66.8) |
| Hypertension, n (%) | 59 (88.1) | 52 (52.5) | 704 (73.4) |
| Cardiovascular Disease, n (%) | 40 (59.7) | 17 (17.2) | 359 (37.4) |
| Type 2 Diabetes, n (%) | 50 (74.6) | 31 (31.3) | 440 (45.9) |
| Chronic kidney disease, n (%) | 42 (62.7) | 13 (13.1) | 300 (31.3) |
| Asthma, n (%) | 18 (26.9) | 23 (23.2) | 294 (30.7) |
| COPD, n (%) | 20 (29.9) | 8 (8.1) | 145 (15.1) |

**Supplementary Table 2.** Breakdown of COVID-19 status across the four studies included in the analysis.

| COVID-19 status | Positive qPCR or serology for SARS-CoV-2 | ICD10 U07 diagnosis or hospitalization | Severe COVID-19 (ventilation or death) | Negative qPCR or serology test for SARS-CoV-2 |  |  |  |  |
| --- | --- | --- | --- | --- | --- | --- | --- | --- |
|  |  |  |  |  | UK Biobank | GHS | PMBB | Total |
| Positive | Yes | Yes | Yes | Yes or No or NA | 382 | 49 | 27 | 458 |
|  | Yes | Yes | No or NA | Yes or No or NA | 1119 | 121 | 40 | 1,280 |
|  | Yes | No or NA | Yes | Yes or No or NA | 0 | 0 | 0 | 0 |
|  | Yes | No or NA | No or NA | Yes or No or NA | 5400 | 664 | 99 | 6,163 |
|  | No or NA | Yes | Yes | No or NA | 122 | 0 | 0 | 122 |
|  | No or NA | Yes | No or NA | No or NA | 77 | 0 | 0 | 77 |
|  | No or NA | No or NA | Yes | No or NA | 0 | 0 | 0 | 0 |
|  | No or NA | Yes | Yes | Yes | 37 | 0 | 0 | 37 |
|  | No or NA | Yes | No or NA | Yes | 111 | 0 | 0 | 111 |
|  | No or NA | No or NA | Yes | Yes | 0 | 0 | 0 | 0 |
|  |  |  |  |  | Total = 7248 | Total = 834 | Total = 166 | Total = 8248 |
| Negative | No or NA | No or NA | No or NA | Yes | 32,867 | 15,574 | 959 | 49,400 |
| Unknown | NA | No or NA | No or NA | NA | 384,068 | 93,718 | 7,779 | 485,565 |
| Total |  |  |  |  | 424,183 | 110,126 | 8,904 | 543,213 |

**Supplementary Table 3.**
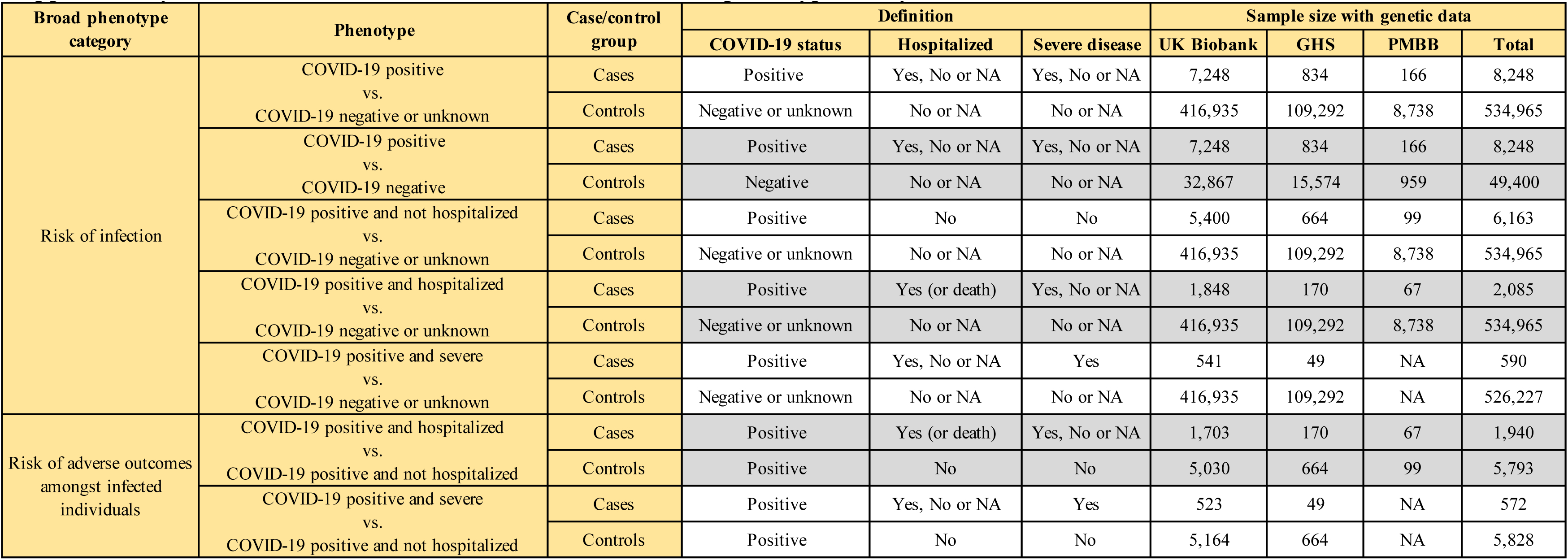
Definitions used for the seven COVID-19 phenotypes analyzed.

**Supplementary Table 4.**
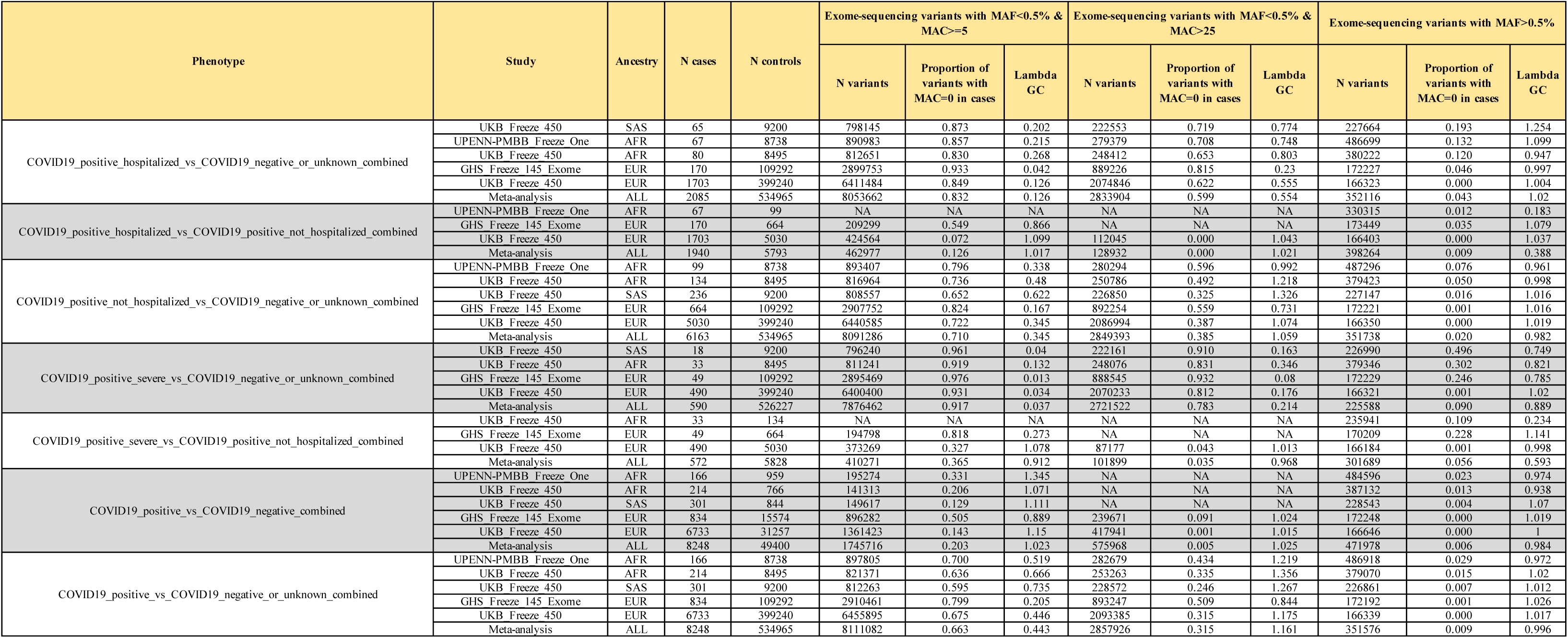
Genomic inflation factor (λ_GC_) observed in the analysis of exome sequence variants for each of the eight phenotypes tested.

**Supplementary Table 5.**
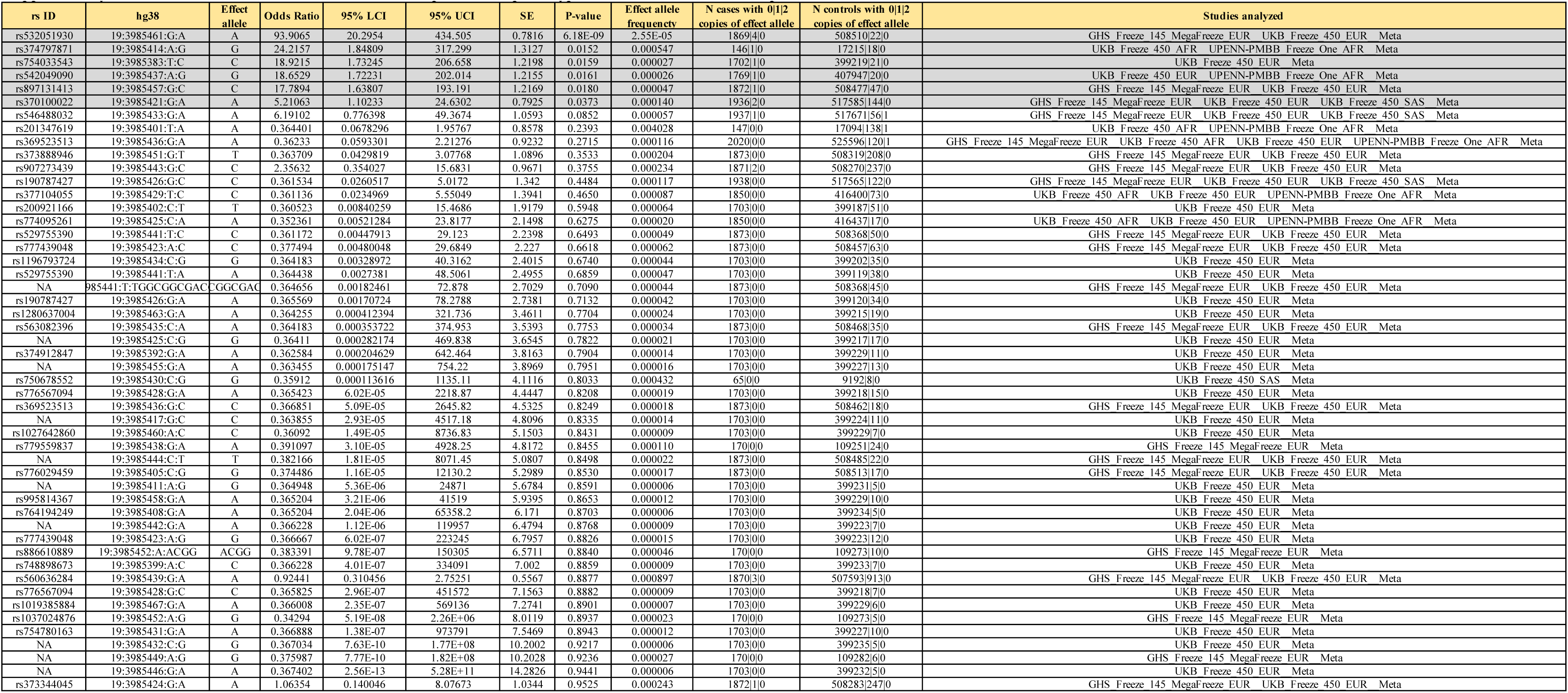
Association between the COVID-19 hospitalization phenotype and 50 rare variants in the promoter of *EEF2*.

**Supplementary Table 6.**
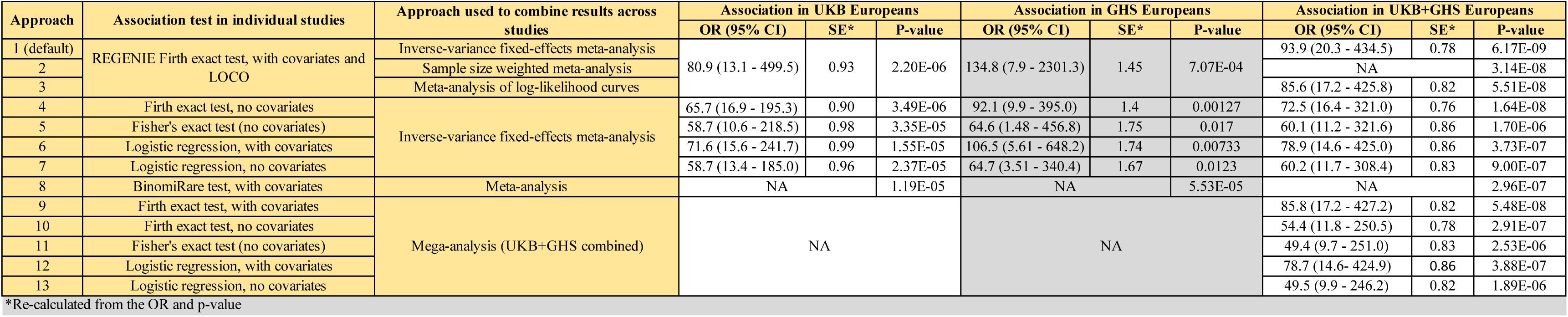
Evidence for association between the COVID-19 hospitalization phenotype and rs532051930 across different association tests.

**Supplementary Table 7.** No carriers of the rare variant rs532051930 in the promoter of EEF2 were observed in an additional 4,122 individuals with COVID-19.

| Study | Reference | N COVID-19 cases |  |  |  |  |  |  |  |  |
| --- | --- | --- | --- | --- | --- | --- | --- | --- | --- | --- |
|  |  | Total | Hospitalized | Severe | Of European | African | Asian | Hispanic | other ancestry | Heterozygote for rs532051930:A |
| GenOMICC | PMID 33307546 | 2969 | 2969 | 2969 | 2110 | 145 | 375 | NA | 339 | 0 |
| Columbia University COVID-19 Biobank | https://www.medrxiv.org/content/10.1101/2020.12.18.20248226v2 | 1152 | 1082 | 479 | 67 | 463 | 33 | 506 | 83 | 0 |
| Biobanque Québec Covid-19 | https://www.medrxiv.org/content/10.1101/2020.12.18.20248226v2 | 220 | 128 | 62 | 154 | 30 | 29 | 7 | 0 | 0 |

**Supplementary Table 8.**
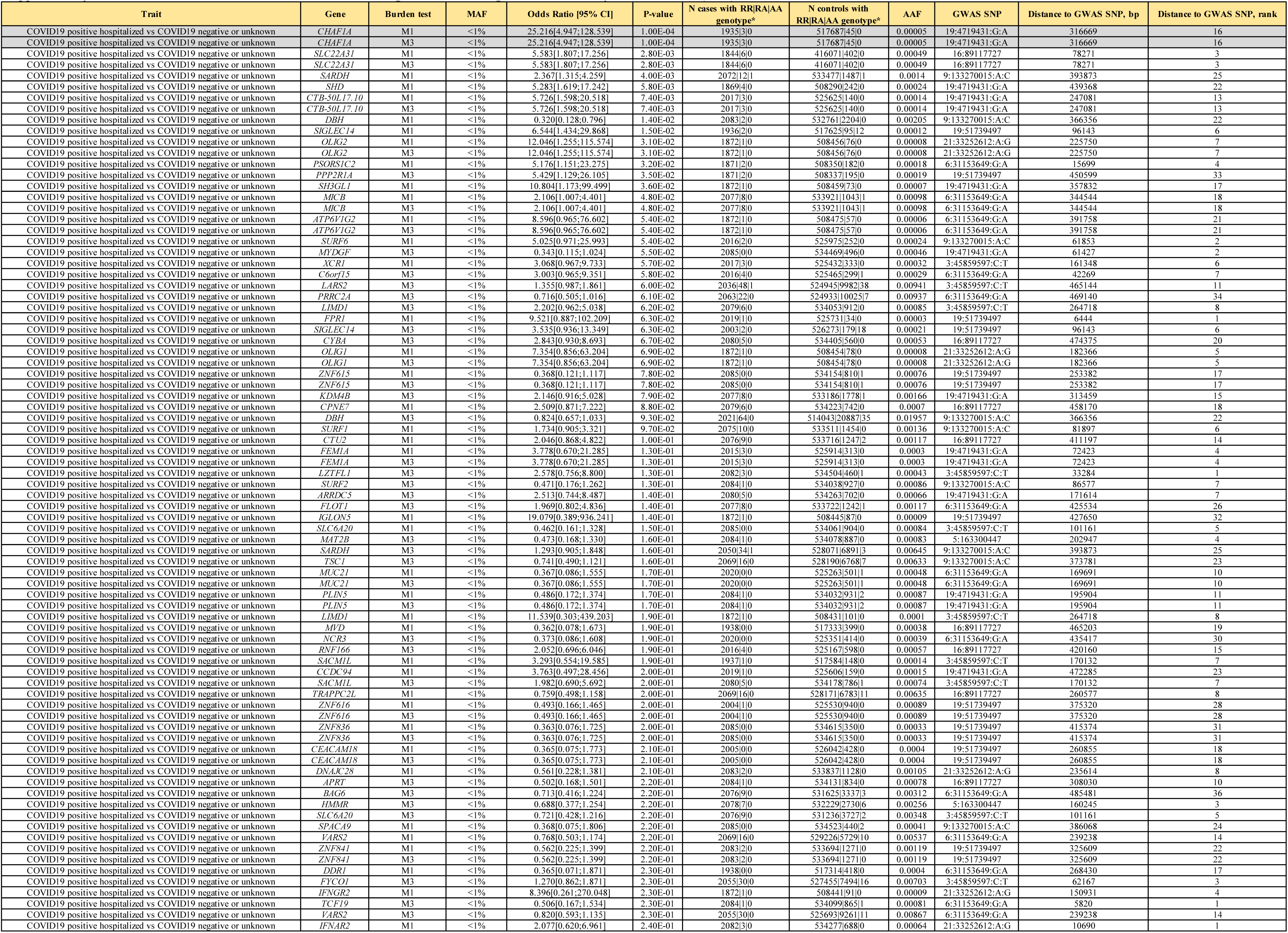
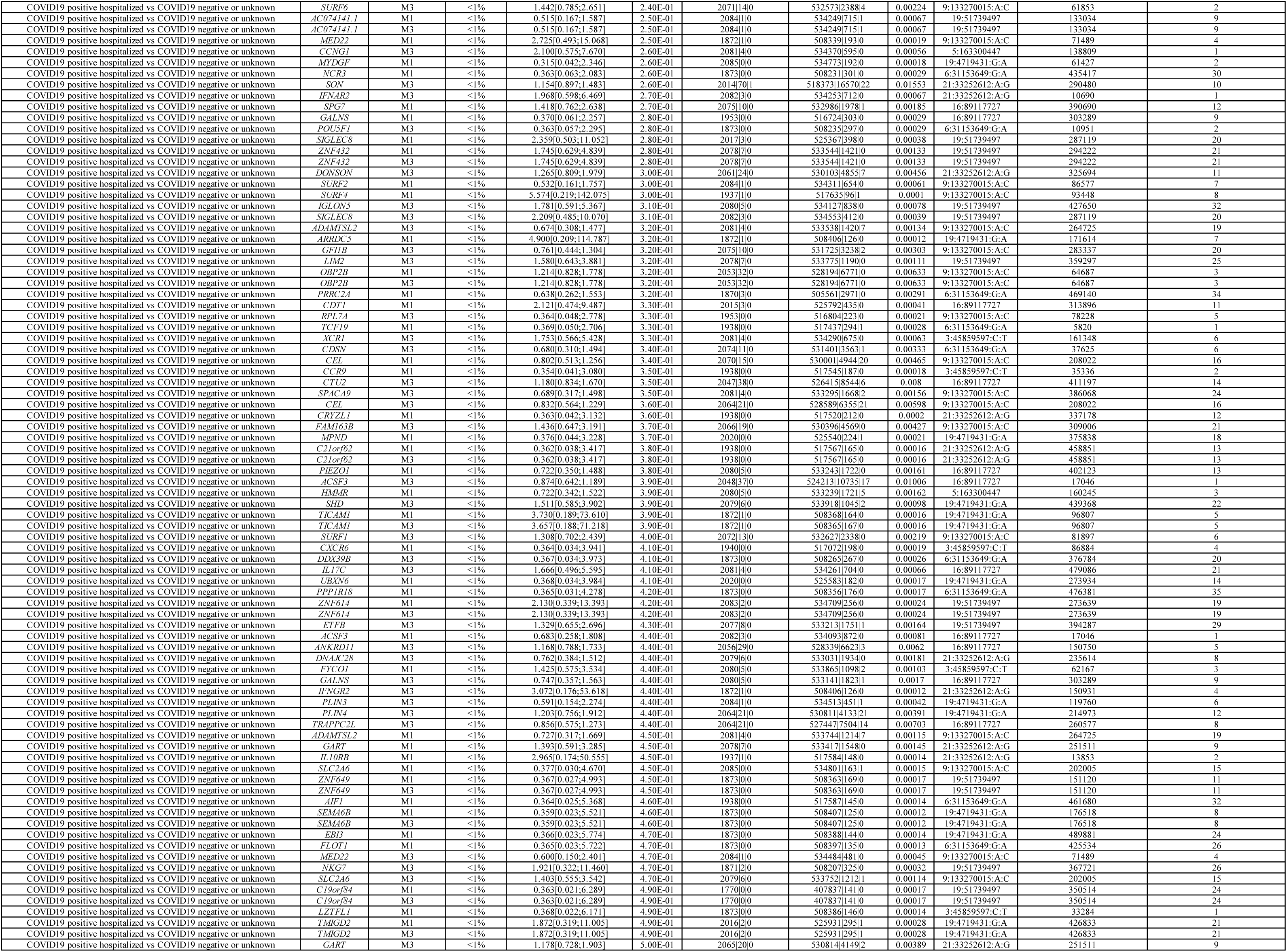
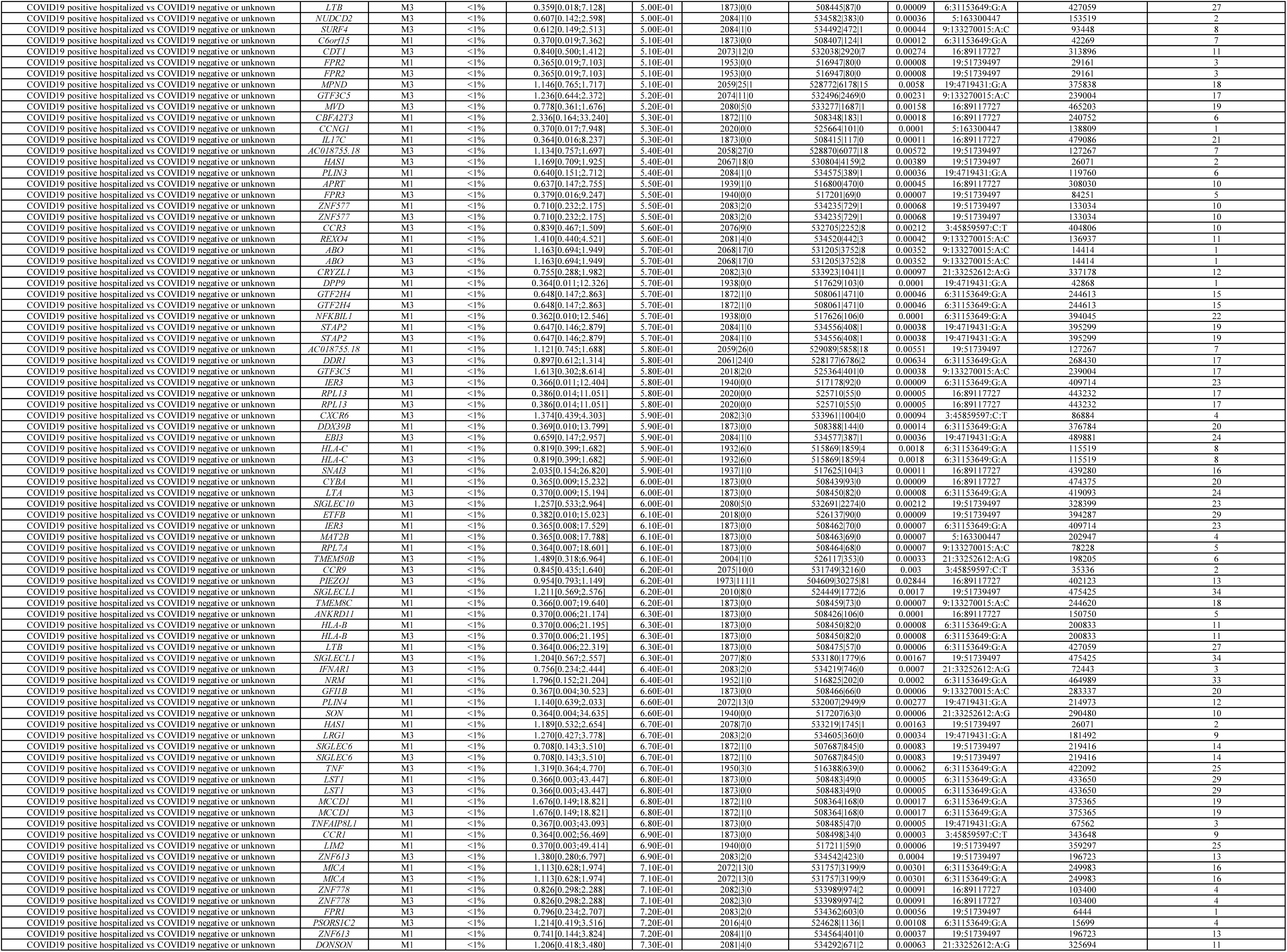
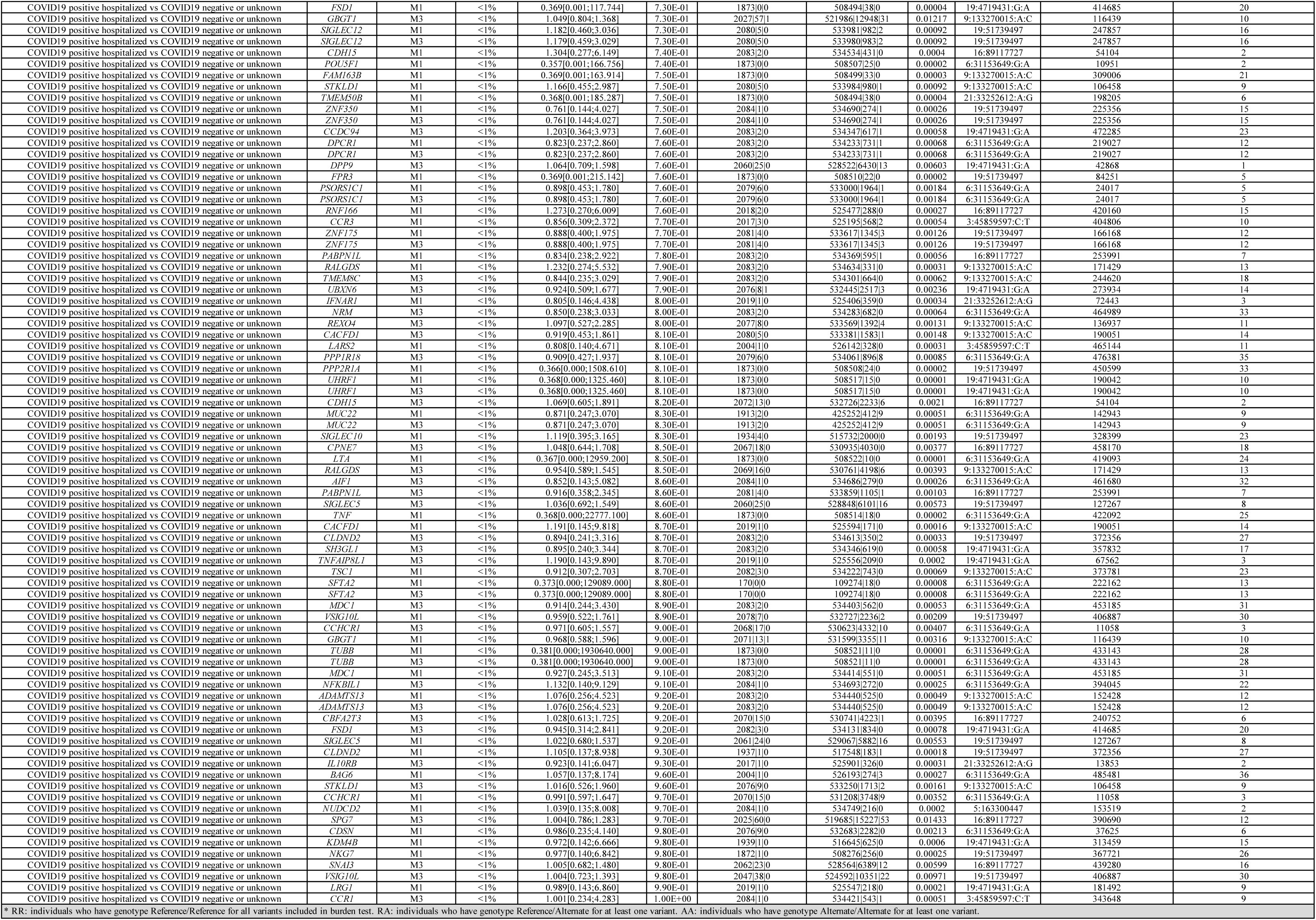
Results from burden association tests for 167 genes located in eight loci described in Horowitz et al. [13].

**Supplementary Table 9.**
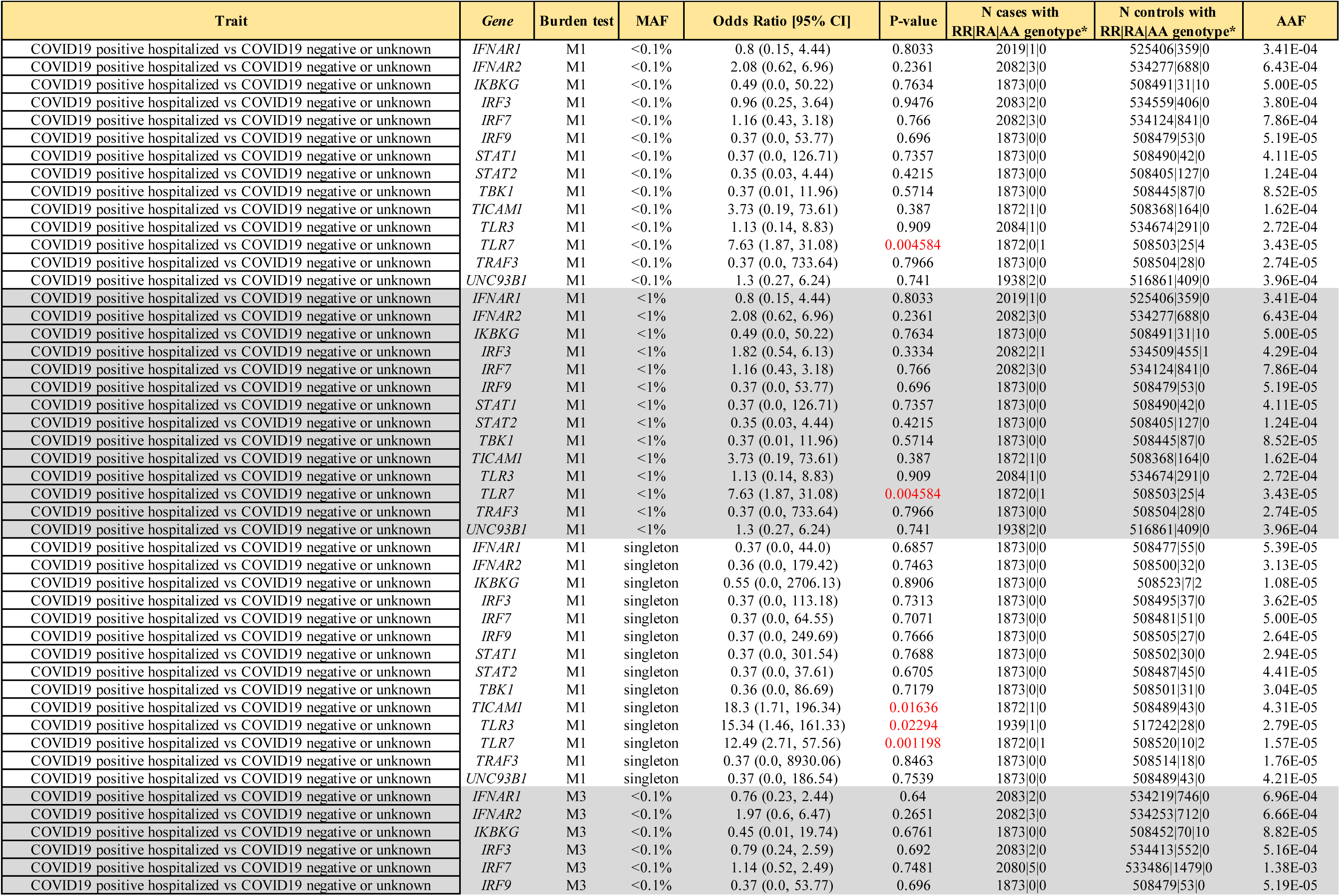
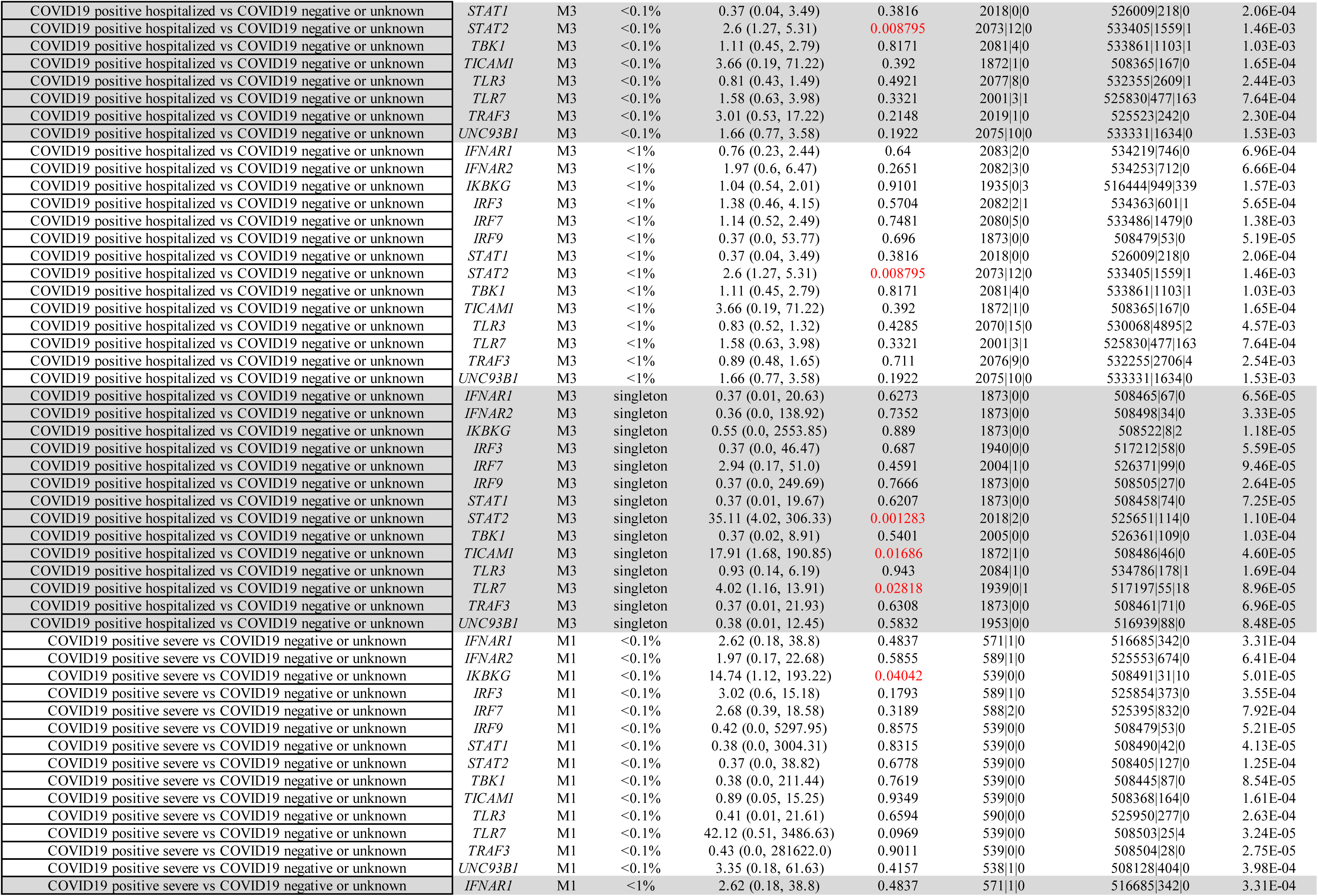
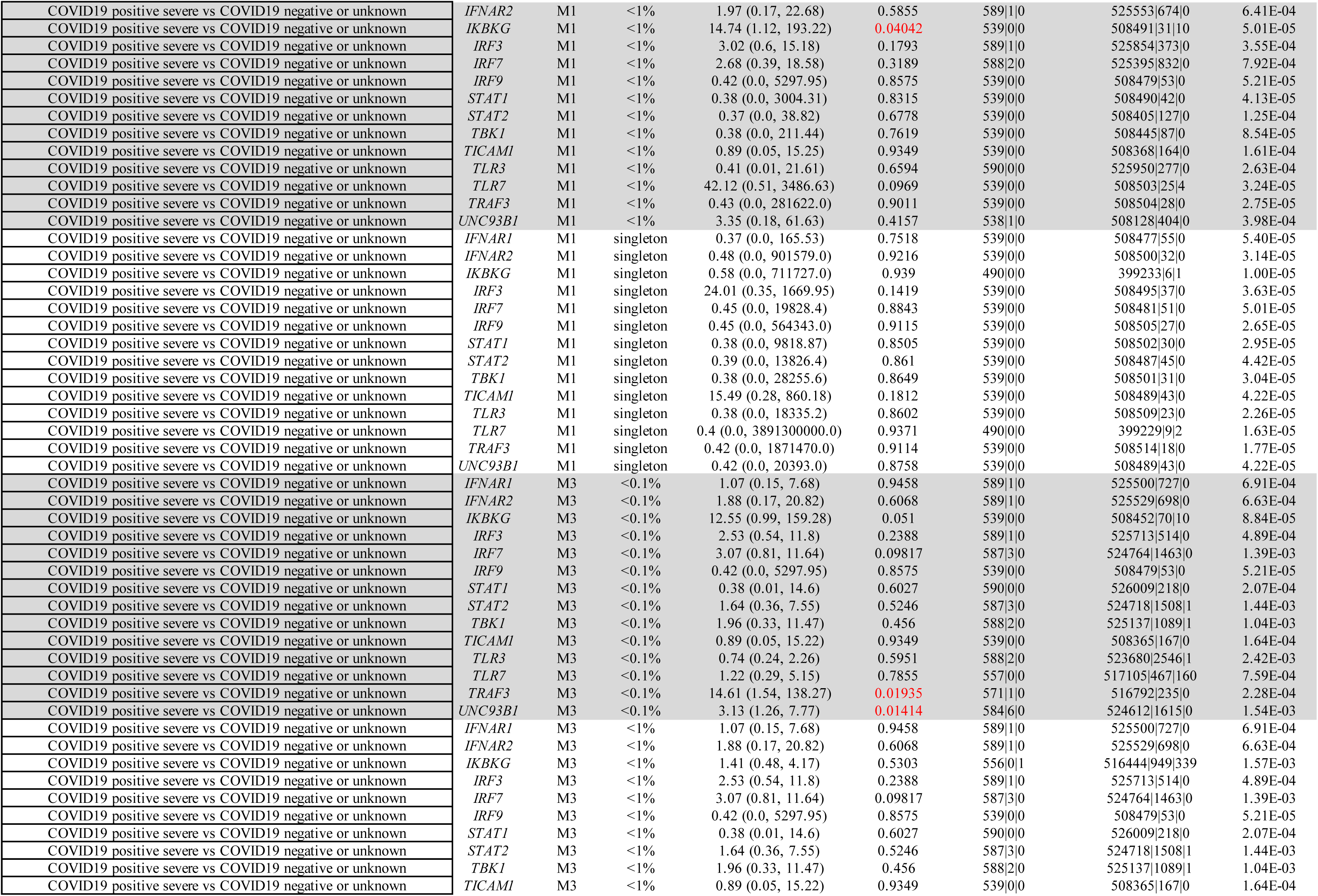
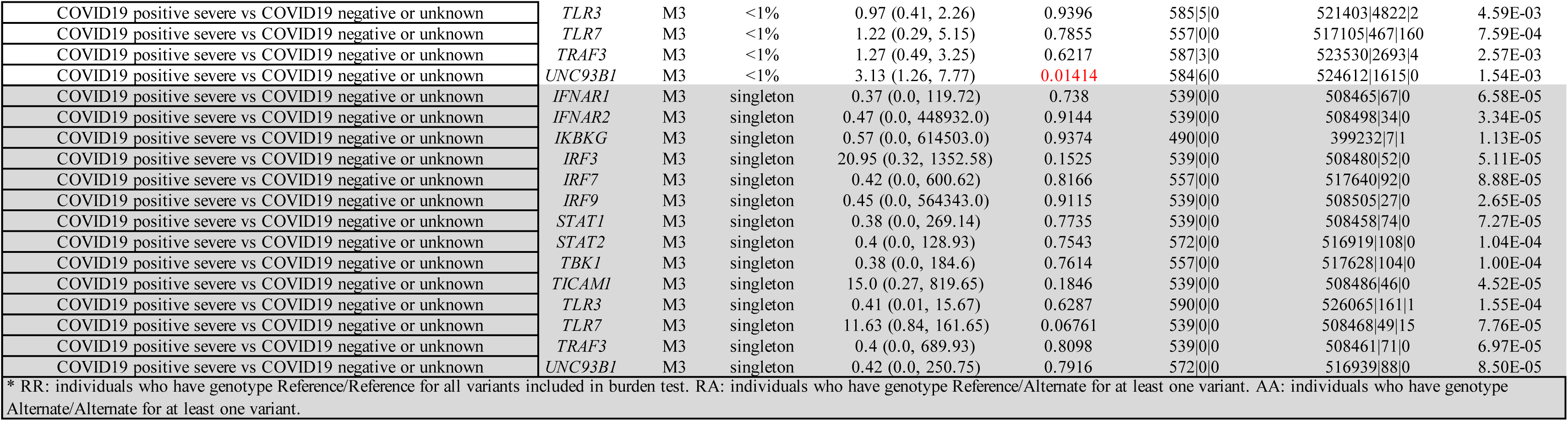
Results from burden association tests for 14 genes related to interferon signaling and recently reported to contain rare (MAF<0.1%), deleterious variants in patients with severe COVID-19.

**Supplementary Table 10.**
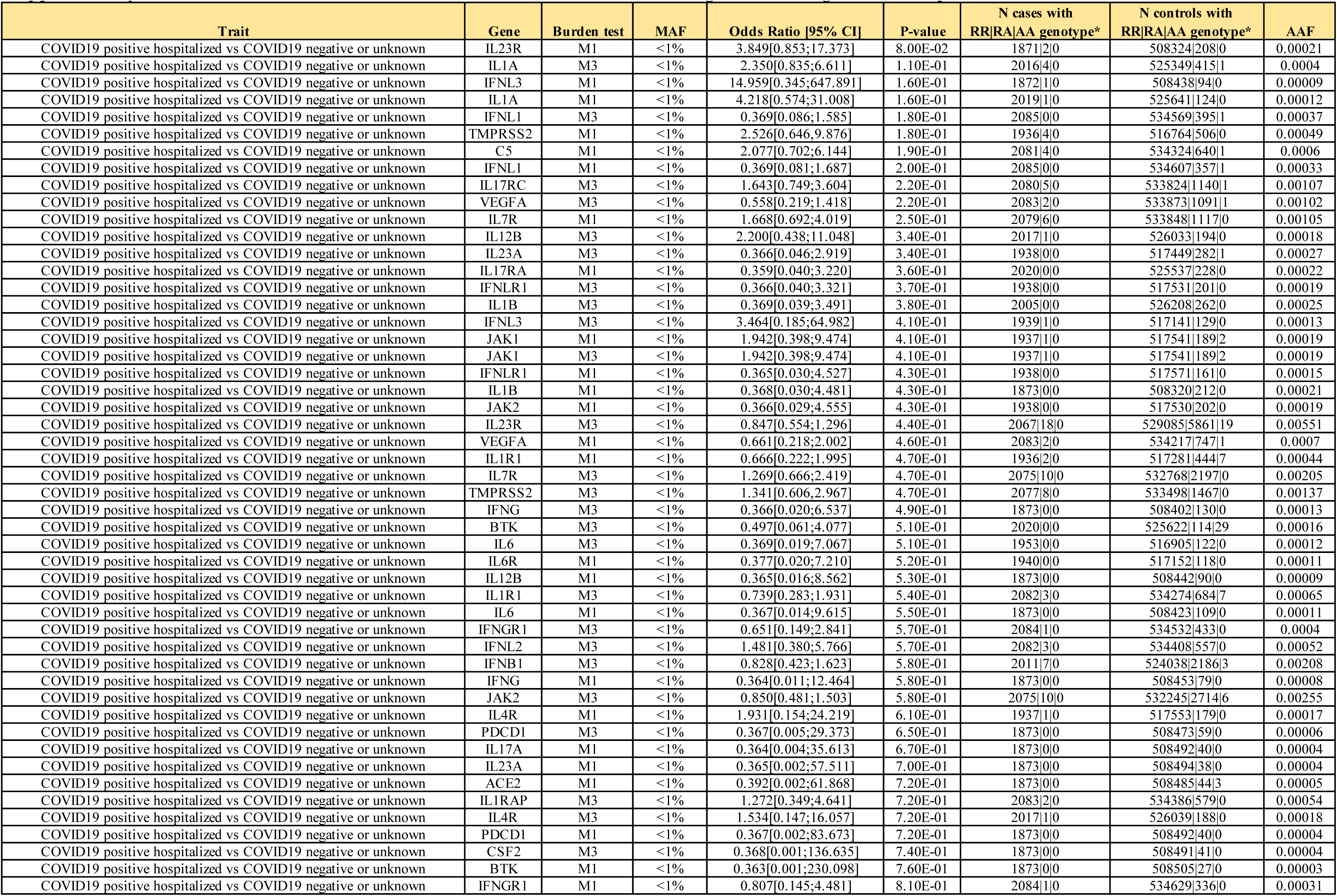
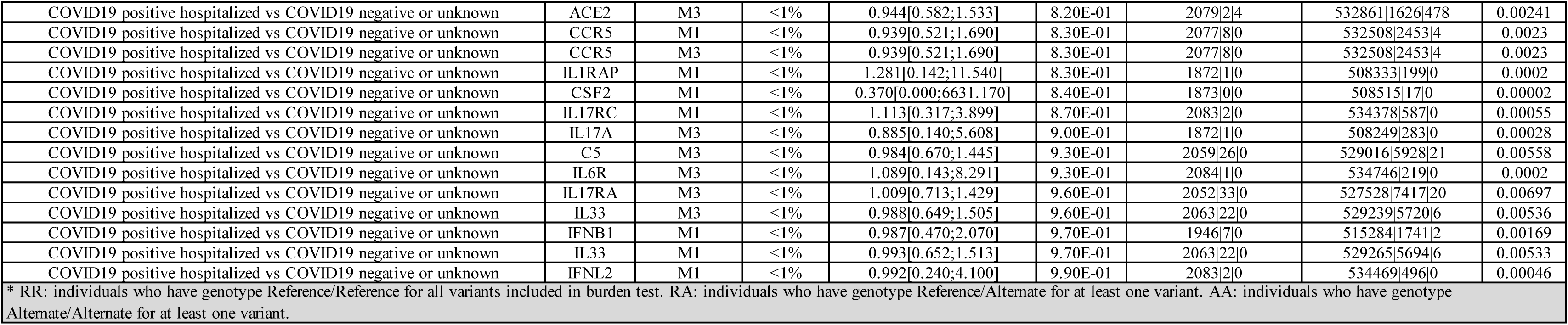
Results from burden association tests for an additional 32 genes that are involved in the etiology of SARS-CoV-2, encode therapeutic targets or have been implicated in other immune or infectious diseases through GWAS.

## SUPPLEMENTARY TEXT

### Regeneron Genetics Center (RGC) Research Team and Contribution Statements

All authors/contributors are listed in alphabetical order.

#### RGC Management and Leadership Team

Goncalo Abecasis, Ph.D., Aris Baras, M.D., Michael Cantor, M.D., Giovanni Coppola, M.D., Aris Economides, Ph.D., Luca A. Lotta, M.D., Ph.D., John D. Overton, Ph.D., Jeffrey G. Reid, Ph.D., Alan Shuldiner, M.D.

Contribution: All authors contributed to securing funding, study design and oversight. All authors reviewed the final version of the manuscript.

#### Sequencing and Lab Operations

Christina Beechert, Caitlin Forsythe, M.S., Erin D. Fuller, Zhenhua Gu, M.S., Michael Lattari, Alexander Lopez, M.S., John D. Overton, Ph.D., Thomas D. Schleicher, M.S., Maria Sotiropoulos Padilla, M.S., Louis Widom, Sarah E. Wolf, M.S., Manasi Pradhan, M.S., Kia Manoochehri, Ricardo H. Ulloa.

Contribution: C.B., C.F., A.L., and J.D.O. performed and are responsible for sample genotyping. C.B, C.F., E.D.F., M.L., M.S.P., L.W., S.E.W., A.L., and J.D.O. performed and are responsible for exome sequencing. T.D.S., Z.G., A.L., and J.D.O. conceived and are responsible for laboratory automation. M.P., K.M., R.U., and J.D.O are responsible for sample tracking and the library information management system.

#### Clinical Informatics

Nilanjana Banerjee, Ph.D., Michael Cantor, M.D. M.A., Dadong Li, Ph.D., Deepika Sharma, MHI

Contribution: All authors contributed to the development and validation of clinical phenotypes used to identify study subjects and (when applicable) controls.

#### Genome Informatics

Xiaodong Bai, Ph.D., Suganthi Balasubramanian, Ph.D., Andrew Blumenfeld, Gisu Eom, Lukas Habegger, Ph.D., Alicia Hawes, B.S., Shareef Khalid, Jeffrey G. Reid, Ph.D., Evan K. Maxwell, Ph.D., William Salerno, Ph.D., Jeffrey C. Staples, Ph.D.

Contribution: X.B., A.H., W.S. and J.G.R. performed and are responsible for analysis needed to produce exome and genotype data. G.E. and J.G.R. provided compute infrastructure development and operational support. S.B., and J.G.R. provide variant and gene annotations and their functional interpretation of variants. E.M., J.S., A.B., L.H., J.G.R. conceived and are responsible for creating, developing, and deploying analysis platforms and computational methods for analyzing genomic data.

#### Analytical Genetics

Gonçalo R. Abecasis, Ph.D., Joshua Backman, Ph.D., Manuel A. Ferreira, Ph.D., Lauren Gurski, Jack A. Kosmicki, Ph.D., Alexander H. Li, Ph.D., Adam E. Locke, Ph.D., Anthony Marcketta, Jonathan Marchini, Ph.D., Joelle Mbatchou, Ph.D., Shane McCarthy, Ph.D., Colm O’Dushlaine, Ph.D., Dylan Sun, Kyoko Watanabe, Ph.D.

Contribution: J.A.K. and M.A.F. performed association analyses and led manuscript writing group. J.B. identified low-quality variants in exome sequence data using machine learning. L.G. and K.W. helped with visualization of association results. A.H.L., A.E.L., A.M. and D.S. prepared the analytical pipelines to perform association analyses. J.M. and J.M. developed and helped deploy REGENIE. S.M. and C.O’D. helped defined COVID-19 phenotypes. G.R.A. supervised all analyses. All authors contributed to and reviewed the final version of the manuscript.

#### Immune, Respiratory, and Infectious Disease Therapeutic Area Genetics

Julie E. Horowitz, PhD.

Contribution: J.E.H. helped defined COVID-19 phenotypes, interpret association results and led the manuscript writing group.

#### Research Program Management

Marcus B. Jones, Ph.D., Michelle LeBlanc, Ph.D., Jason Mighty, Ph.D., Lyndon J. Mitnaul, Ph.D.

Contribution: All authors contributed to the management and coordination of all research activities, planning and execution. All authors contributed to the review process for the final version of the manuscript.

### UK Biobank Exome Sequencing Consortium Research Team

#### ^1^Bristol Myers Squibb

Oleg Moiseyenko, Carlos Rios, Saurabh Saha

#### ^2^Regeneron Pharmaceuticals Inc

Listed in pages 38 to 40.

#### ^3^Biogen Inc

Sally John, Chia-Yen Chen, David Sexton, Paola G. Bronson, Christopher D. Whelan, Varant Kupelian, Eric Marshall, Timothy Swan, Susan Eaton, Jimmy Z. Liu, Stephanie Loomis, Megan Jensen, Saranya Duraisamy, Ellen A. Tsai, Heiko Runz

#### ^4^Alnylam Pharmaceuticals

Aimee M. Deaton, Margaret M. Parker, Lucas D. Ward, Alexander O. Flynn-Carroll, Greg Hinkle, Paul Nioi

#### ^5^AstraZeneca

Olympe Chazara, Sri VV. Deevi, Xiao Jiang, Amanda O’Neill, Slavé Petrovski, Katherine Smith, Quanli Wang

#### ^6^Takeda California Inc

Jason Tetrault, Dorothee Diogo, Aldo Cordova Palomera, Emily Wong, Rajesh Mikkilineni, David Merberg, Sunita Badola, Erin N. Smith, Sandor Szalma

#### ^7^Pfizer, Inc

Yi-Pin Lai, Xing Chen, Xinli Hu, Melissa R. Miller

#### ^8^Abbvie

Xiuwen Zheng, Bridget Riley-Gillis, Jason Grundstad, Sahar Esmaeeli, Jeff Waring, J. Wade Davis

^1^Bristol Myers Squibb, Route 206 and Province Line Road, Princeton, NJ 08543, USA

^2^Regeneron Pharmaceuticals Inc., 777 Old Saw Mill River Road, Tarrytown, New York 10591, USA

^3^Biogen Inc., 225 Binney Street, Cambridge, MA 02139, USA

^4^Alnylam Pharmaceuticals, 675 West Kendall St, Cambridge, MA 02142, USA

^5^AstraZeneca Centre for Genomics Research, Discovery Sciences, BioPharmaceuticals R&D, Cambridge, UK

^6^Takeda California Inc., 9625 Towne Centre Dr, San Diego, CA 92121, USA

^7^ Pfizer, Inc., 1 Portland Street, Cambridge MA 02139, USA

^8^ AbbVie, Inc., 1 N. Waukegan Rd, North Chicago, IL 60064, USA

### GenOMICC Consortium

Sara Clohisey^1^, Fiona Griffiths^1^, James Furniss^1^, James Furniss^1^, Trevor Paterson^1^, Tony Wackett^1^, Ruth Armstrong^1^, Wilna Oosthuyzen^1^, Nick Parkinson^1^, Max Head Fourman^1^, Andrew Law^1^, Veronique Vitart^2^, Lucija Klaric^2^, Anne Richmond^2^, Chris P. Ponting^2^, Andrew D. Bretherick^2^, Charles Hinds^3^, Timothy Walsh^4^, Sean Keating^4^, Clark D Russell^1,5^, Malcolm G. Semple^6,7^, Kathy Rowan^8^, Elvina Gountouna^9^, Nicola Wrobel^10^, Lee Murphy^10^, Angie Fawkes^10^, Richard Clark^10^, Audrey Coutts^10^, Lorna Donnelly^10^, Tammy Gilchrist^10^, Katarzyna Hafezi^10^, Louise Macgillivray^10^, Alan Maclean^10^, Sarah McCafferty^10^, Kirstie Morrice^10^, Angie Fawkes^10^, Julian Knight^11^, Charlotte Summers^12^, Manu Shankar-Hari^13,14^, Peter Horby^15^, Alistair Nichol^16,17,18^, David Maslove^19^, Lowell Ling^20^, Danny McAuley^21,22^, Hugh Montgomery^23^, Peter J.M. Openshaw^24,25^.

^1^Roslin Institute, University of Edinburgh, Easter Bush, Edinburgh, EH25 9RG, UK

^2^MRC Human Genetics Unit, Institute of Genetics and Molecular Medicine, University of Edinburgh, Western General Hospital, Crewe Road, Edinburgh, EH4 2XU, UK

^3^William Harvey Research Institute, Barts and the London School of Medicine and Dentistry, Queen Mary University of London, London EC1M 6BQ, UK

^4^Intensive Care Unit, Royal Infirmary of Edinburgh, 54 Little France Drive, Edinburgh, EH16 5SA, UK

^5^Centre for Inflammation Research, The Queen’s Medical Research Institute, University of Edinburgh, 47 Little France Crescent, Edinburgh, UK

^6^NIHR Health Protection Research Unit for Emerging and Zoonotic Infections, Institute of Infection, Veterinary and Ecological Sciences University of Liverpool, Liverpool, L69 7BE, UK

^7^Respiratory Medicine, Alder Hey Children’s Hospital, Institute in The Park, University of Liverpool, Alder Hey Children’s Hospital, Liverpool, UK

^8^Intensive Care National Audit & Research Centre, London, UK

^9^Centre for Genomic and Experimental Medicine, Institute of Genetics and Molecular Medicine, University of Edinburgh, Western General Hospital, Crewe Road, Edinburgh, EH4 2XU, UK

^10^Edinburgh Clinical Research Facility, Western General Hospital, University of Edinburgh, EH4 2XU, UK

^11^Wellcome Centre for Human Genetics, University of Oxford, Oxford, UK

^12^Department of Medicine, University of Cambridge, Cambridge, UK

^13^Department of Intensive Care Medicine, Guy’s and St. Thomas NHS Foundation Trust, London, UK

^14^School of Immunology and Microbial Sciences, King’s College London, UK

^15^Centre for Tropical Medicine and Global Health, Nuffield Department of Medicine, University of Oxford, Old Road Campus, Roosevelt Drive, Oxford, OX3 7FZ, UK

^16^Clinical Research Centre at St Vincent’s University Hospital, University College Dublin, Dublin, Ireland

^17^Australian and New Zealand Intensive Care Research Centre, Monash University, Melbourne, Australia

^18^Intensive Care Unit, Alfred Hospital, Melbourne, Australia

^19^Department of Critical Care Medicine, Queen’s University and Kingston Health Sciences Centre, Kingston, ON, Canada

^20^Department of Anaesthesia and Intensive Care, The Chinese University of Hong Kong, Prince of Wales Hospital, Hong Kong, China

^21^Wellcome-Wolfson Institute for Experimental Medicine, Queen’s University Belfast, Belfast, Northern Ireland, UK

^22^Department of Intensive Care Medicine, Royal Victoria Hospital, Belfast, Northern Ireland, UK

^23^UCL Centre for Human Health and Performance, London, W1T 7HA, UK

^24^National Heart and Lung Institute, Imperial College London, London, UK

^25^Imperial College Healthcare NHS Trust: London, London, UK 848

